# Genomic Surveillance Identifies SARS-CoV-2 Transmission Patterns in Local University Populations, Wisconsin, USA, 2020–2022

**DOI:** 10.1101/2022.07.06.22277014

**Authors:** Arunachalam Ramaiah, Manjeet Khubbar, Amy Bauer, Katherine Akinyemi, Joshua Weiner, Sanjib Bhattacharyya

## Abstract

Novel variants of severe acute respiratory syndrome coronavirus 2 (SARS-CoV-2) continue to emerge as the current coronavirus disease 2019 (COVID-19) pandemic extends into its third year. Understanding SARS-CoV-2 circulation in university populations is vital for effective interventions in a higher education setting that will inform pubic health policy during pandemics. In this study, we performed whole-genome sequencing of 537 of 1,717 SARS-CoV-2 positive nasopharyngeal/nasal swab samples collected for nearly 20 months from the two university populations in Wisconsin, United States. We observed that the viral sequences were distributed into 57 lineages/sub-lineages belonging to 15 clades of which the majority were from 21K (Omicron, 36.13%) and 21J (Delta, 30.91%). Nearly 40% (213) of the sequences were Omicron of which BA.1 and its eight descendent lineages account for 91%, while the remaining belong to BA.2 and its six descendent lineages. The independent analysis of these two universities sequences revealed significant differences in circulating the SARS-CoV-2 variants. The genome-based analysis of closely-related strains along with phylogenetic clusters had identified that potential virus transmission occurred within as well as between universities, and between the university and local community. Although this study improves our understanding of distinct transmission patterns of circulating variants in local universities, expanding the genomic surveillance capacity will aid local jurisdictions in identifying emerging SARS-CoV-2 variants like BA.4 and BA.5, and improve data-driven public health mitigation and policy efforts.

## Introduction

Severe Acute Respiratory Syndrome Coronavirus 2 (SARS-CoV-2), the causative agent of the Coronavirus disease 2019 (COVID-19) pandemic, is a positive-sense, single-stranded RNA virus with approximately 30 kilobases of genome. SARS-CoV-2 was first detected in Wuhan, China in 2019 and rapidly spread across the globe prompting a massive human health crisis [1,2]. SARS-CoV-2 has infected over 542 million people and claimed over 6.3 million lives at time of writing. The pandemic rages unabated despite nearly 12 billion vaccine doses having been administered (WHO Coronavirus (COVID-19) Dashboard).

Genomic surveillance of SARS-CoV-2 provides key insights into virus evolution, transmission dynamics, and circulation of variants in a diverse spectrum of communities from different geographical areas [3]. The majority of COVID-19 cases in Wisconsin were reported from correctional and long-term care facilities, colleges and universities [4], and unvaccinated people. COVID-19 transmission in institutions of higher education across the United States were reported as a result of resuming in-person instruction of large groups, extensive social gatherings, high-density clustering with on-campus housing, and significant number of off-campus student population [4-7]. While university students are less likely to develop severe COVID-19, there is still a possibility that outbreaks in the university could spread to more vulnerable populations in the local community [8,9]. Therefore, it is vital to identify circulating virus variants and possible chains of cross-transmission among university and local community as this will aid local jurisdictions in identifying emerging variants and improving public health mitigation and policy efforts. As part of this effort, the City of Milwaukee Health Department Laboratory (MHDL) has partnered with the health clinics of two Milwaukee, Wisconsin-based universities (Marquette University, MU; University of Wisconsin-Milwaukee, UWM) since September 2020, and has contributed to the testing of 1,717 nasopharyngeal/nasal swab samples of students and staff providing diagnostic testing and genomic surveillance for SARS-CoV-2. We selected a group of 537 SARS-CoV-2 positive samples collected between September 2020 and April 2022 for whole genome sequencing at the MHDL using the MiSeq (Illumina, San Diego, CA), MinION (Nanopore, San Francisco, CA) and Clear Dx™ (Clear Labs, San Carlos, CA) sequencing platforms. Overall, this genomic surveillance and bioinformatic data analysis identified the circulating lineages and descendent lineages, and the transmission dynamics of SARS-CoV-2 within as well as between universities, and between the university and the local community, highlighting the importance of MHDL molecular surveillance efforts in identification of outbreak clusters on these universities’ campuses to monitor and track suspected cases.

## Methods

### Samples

The City of Milwaukee Health Department Laboratory has partnered with the student health clinics of University of Wisconsin-Milwaukee and Marquette University, Milwaukee, Wisconsin and has tested 1,717 human specimens (nasopharyngeal/nasal swab), which were collected for SARS-CoV-2 diagnosis between September 2020 and April 2022 according to CDC Interim Guidelines for Collecting and Handling of Clinical Specimens for COVID-19 Testing and transported to MHDL in a timely manner [10]. The MHDL Microbiology Requisition form was used to get all patient demographic information, which was entered into MHDL’s laboratory information system at the initial time of the SARS-CoV-2 diagnostic testing and was later retrieved in spreadsheet format for further analysis.

### SARS-CoV-2 RNA extraction

All specimens or nucleic acid specimens used in this study were obtained under Milwaukee Health Department (MHD) Institutional Biosafety Committee (IBC) guidelines, and per MHD Institutional Review Board (IRB) and Ethics Committee approval. MHD IRB and ethics committee determined the laboratory analysis and genomic epidemiological activities were for non-research public health surveillance and had no ethical concerns. Nucleic acid extractions were performed using FDA EUA approved automated extraction platforms such as Maxwell rapid sample concentrator (RSC) platform (Promega, Madison, WI), NucliSens easyMAG, EMAG (bioMérieux, Boxtel, The Netherlands), EZ1 Advanced XL (Qiagen, Valencia, CA), and KingFisher Flex platforms (Thermo Fisher Scientific, Waltham, MA) according to manufacturer’s instructions and extract stored at -80°C until use [11,12]. Real-time RT-PCR setup was performed using 5 µL of extract and EUA-approved CDC 2019-Novel Coronavirus (2019-nCoV) Real-Time Reverse Transcriptase (RT)-PCR Diagnostic Panel or Applied Biosystems TaqPath™ COVID-19 Combo Kit or on Cepheid’s GeneXpert Dx instrument using Xpert Xpress SARS-CoV-2 assay. Nucleic acid extraction controls for each batch of clinical specimens were used for checking quality. Various PCR panels have been used throughout the pandemic as tests and instruments became available and modifications were made to existing panels.

### SARS-CoV-2 sequencing and data analysis

Representative samples (n=562) used for genomic sequencing were from those two university populations specimens that tested positive for SARS-CoV-2 using diagnostic PCR assays authorized for use by the Food and Drug Administration (FDA) under an Emergency Use Authorization (EUA) [11,12]. Sample selection for sequencing varied quite a bit. There were different QC metrics depending on the instrument being used. During the early stage of sequencing, the recommended QC metrics were also constantly being adjusted depending on the variant being sequenced and the progression of the pandemic. Samples were then checked after sequencing and only curated if they passed post-run quality control metrics. Samples with <28 ct value were successfully sequenced using the ARTIC protocol and the Illumina DNA Prep library kit on MiSeq instrument [13]. Data generated using the Integrated DNA technologies (IDTs) ARTIC V3 or V4 primer panel on MiSeq was analyzed using the Illumina BaseSpace™ DRAGEN COVID Lineage App (v3.5.4 to v3.5.9), which uses a customized version of the DRAGEN DNA pipeline to perform Kmer-based detection of SARS-CoV-2. This App aligns reads to a reference genome, calls variants, and generates a consensus genome sequence. Lineage/clade assignments are also confirmed using NextClade and Pangolin COVID-19 Lineage Assigner by uploading obtained FASTA files.

SARS-CoV-2 whole-genome sequencing was also performed using the Clear Labs ClearDx™, a fully automated, next-generation sequencing platform. The fully automated test goes from extracted RNA from SARS-CoV-2 positive clinical respiratory specimens to generation of compressed FASTQ and FASTA files including sequencing coverage and assembly coverage with minimal human intervention. The Clear Dx(tm) system uses a Hamilton STAR robotic platform for liquid handling, Hamilton thermal cyclers, a barcode reader, magnet block, and two MinION nanopore sequencers from Oxford Nanopore Technologies. Total nucleic acid was extracted from SARS-CoV-2 positive clinical specimens and amplified using ARTIC V3 or MIDNIGHT primer pools. For each run, 30 known SARS-CoV-2 RNA extracts from clinical samples that met quality control requirements of < 30 Ct value were tested as per manufacturer guidelines. Synthetic SARS-CoV-2 RNA controls procured from Twist Biosciences were included as a positive control (Twist Biosciences, San Francisco, CA). Stock solution of synthetic SARS-CoV-2 RNA controls was serially diluted from 10,000 copies/μL to 1 copy/μL before use. Upon completion of sequencing, FASTA / FASTQ files and quality metrics for each sample were available for download for downstream analysis.

For Nanopore MinION and Clear Labs reads, BIP-WV6, BIP-WV7, ARTIC Pipeline, and Medaka via artic 1.2.1 were used to assemble the SARS-CoV-2 genome. Consensus sequences generated and related metadata for clinical samples are shared publicly on Global Initiative on Sharing All Influenza Data (GISAID; https://www.gisaid.org/), the principal repository for SARS-CoV-2 genetic information.

### Clade and lineage assignation and phylogenetic analysis

The clade and lineage assignment for each of 537 SARS-CoV-2 isolates were performed using Nextclade v1.14.1 [14], Pangolin COVID-19 Lineage Assigner [15], and Ultrafast Sample placement on Existing tRee (UShER) [16]. The closely related genomes and metadata in entire EpiCoV database for the representative circulating Omicron strains in the university population were identified using Audacity*Instant* searches in GISAID. For the phylogenetic analysis, all 537 SARS-CoV-2 along with a reference Wuhan-Hu-1/2019 (GenBank # MN908947) genome sequences were aligned using MAFFT v.7.505 [17] and subsequently these aligned sequences were used to identify GTR+F+R2 as a best fit model based on the Bayesian Information Criteria using ModelFinder [18]. The phylogenetic tree of SARS-CoV-2 was estimated using maximum likelihood (ML) method with 1000 bootstrap replicates in IQ-TREE multi core version 2.0.3 [19]. The phylogenetic tree was annotated with metadata in Interactive Tree Of Life (iTOL) [20]. The figures were generated using Prism GraphPad v8.4.3 (www.graphpad.com).

## Results and discussion

In total, 1,717 nasopharyngeal/nasal swab samples of various students and staff were received at MHDL from two universities for SARS-CoV-2 diagnosis between September 2020 and April 2022. Among 1,717 samples, we selected a group of 562 samples for sequencing, of which 25 genome sequences showing >10% missing nucleotides and less than 100x coverage with poor-quality results were excluded. The remaining 537 (95.6%) sequences (Supplemental data) were analyzed using Illumina DRAGEN COVID lineage app available on BaseSpace (https://basespace.illumina.com/), Clear Labs (https://www.clearlabs.com/), Pangolin COVID-19 Lineage Assigner (https://pangolin.cog-uk.io/), or Nextclade (https://clades.nextstrain.org/) software platforms. Our findings showed that the university populations sequences were distributed into 15 clades, in which >80% of the sequences were derived from three clades: 21K (Omicron) (194, 36.13%), 21J (Delta) (166, 30.91%), and 20G (B.1.2) (74, 13.78%) (Figure 1A and 1B). While this data includes most of the variants of concern (VOC) and variants of interest (VOI), there were no sequences detected for clades 20H (Beta), 20J (Gamma), 21G (Lambda), 22A (Omicron, BA.4) and 22B (Omicron, BA.5). At the time of this study period, 57 lineages/sub-lineages belonging to 15 clades of SARS-CoV-2 were circulating in the two universities (Figure S1). The maximum of 19 and 9 lineages/sub-lineage types belonging to 21J/Delta and 21K/Omicron clades, respectively, were identified in these samples. Nearly 40% (213) of the 537 sequences were Omicron, in which BA.1 and its eight descendent lineages account for 91%, while the remaining belong to BA.2 and its six descendent lineages. Among these sub-lineages BA.1.1 (88) and BA.2.12.1 (4) are most commonly identified in BA.1 and BA.2 lineages respectively (Figure 1B; Figure S1), highlighting that these lineages should be monitored closely for their clinical outcomes and their impact on diagnostics. BA.1 and BA.2.10 were among the 16 Omicron lineage/sub-lineages studied in the university samples detected first in December 2021 and January 2022, which was earlier than other variants identified.

**Figure 1.**
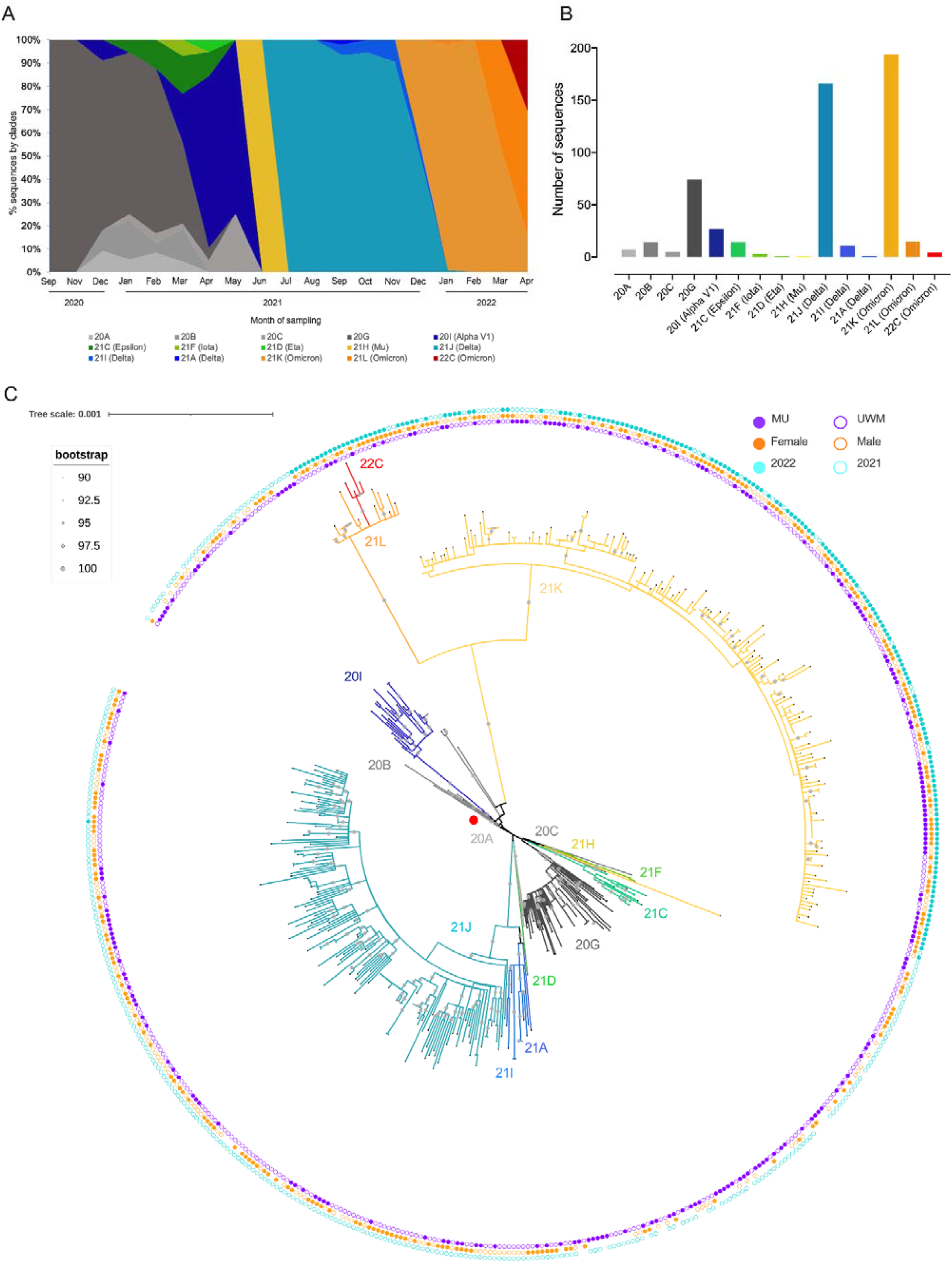
The chronologic distributions and phylogenetic analysis of SARS-CoV-2 sequences from two universities in Milwaukee, Wisconsin from samples collected during September, 2020-April, 2022. **A**) Chronologic distribution of SARS-CoV-2 genomic variants for 20 months in the Milwaukee university’s populations. The pangolin clades for 537 genome sequences of SARS-CoV-2 were identified in Nextclade and classified based on the sampling date from September 2020-April 2022. Clades are color coded following the naming convention and branching colors in Nextclade. **B**) Cumulative number of sequences segregated based on the Nextclade classification. The greater number of sequences belonged to 21K (Omicron) and 21J (Delta) clades. **C**) The schematic representation of the phylogenetic relationship of all 537 SARS-CoV-2 genomes sequenced from clinical specimens of the two university’s populations in Milwaukee, Wisconsin. These sequences aligned against the reference strain Wuhan-Hu-1/2019 (GenBank # MN908947; red circle) before constructing the tree. The maximum likelihood circular tree showed three major clades, one consisting of two major clusters, including 20A, 20C, 20G, 21C, 21F, and 21H, a second one consisting of two clusters, including 21I and 21J, and a third one consisting of 20B, 20I, 21L and 21K clade strains, following the naming convention and branching colors as in Panel A. As almost all the branches are supported by >70 bootstrap values, we display values if they are ≥90% supported (gray circles in the middle of the branches). Most of the nodes in the tree were formed with 100% bootstrap supports (large gray circles in the middle of the branches), confirming that the split of the branches supported with high confidence. Each sequence in the tree is highlighted by a black tip. The distance corresponding to substitution per site is indicated by a scare bar. The metadata are visualized as circles exterior to the branches in the tree. From the inner to the outer concentric circle of the branches: Circle 1–3 are indicated samples that are specific to each of two universities, gender and year of sample collection, respectively. In circle 1 (purple), the filled and outlined circle symbols exemplify that the clinical samples used for sequencing originated from Marquette University (MU) and University of Wisconsin-Milwaukee (UWM), respectively. In circle 2 (orange), gender female (filled circle symbol)) and male (outlined) shown, while, the absence of symbol indicated that no gender data available for the given samples. While the filled and outlined circle symbols in the circle 3 (cyan) indicated the years 2022 and 2021, respectively, the absence of symbol indicated 2020, in which the samples were collected from the patients.

To identify the significant differences in the distribution of SARS-CoV-2 variants between these two universities, we analyzed two datasets individually. This analysis revealed that there were no 21F (Iota), 21D (Eta), 21H (Mu), and 21A (Delta) variants detected in the MU samples. In contrast, there were a few identified in UWM for these variants (Figure S2), suggesting that having more off-campus university populations might be the potential cause for identifying more variants at UWM. Delta (21J) and Omicron (21K) VOCs were detected in both universities in July 2021 and December 2021, respectively, indicating that common patterns appeared in the introduction of variants to these universities’ populations. The Delta variant was first identified in the universities in July 2021, and increased in incidence up to November 2021. During this period, no other clades were detected, indicating that the Delta variant accounted for 100% of circulating SARS-CoV-2 variants. Delta then decreased from December 2021 to January 2022 (21%), corresponding to the rapid emergence and detection of the first Omicron variant (21K, BA.1.20) on December 13, 2021 in both university’s samples (Figure 1; Figure S2). Since February 2022, the Omicron variant has accounted for 100% of the infections.

To shed light on phylogenetic relationships of the 537 SARS-CoV-2 genomes, we constructed the maximum likelihood tree that consists of three major phylogenetic clades, which were further grouped into many clusters (Figure 1C). SARS-CoV-2 from clusters 21J (Delta), and 21L, 21K and 22C (Omicrons) were phylogenetically more diverged from the reference strain. The Omicron variants reported were closely related to the Alpha variant, confirming that evolutionary pressures caused the novel Omicron variants to differentiate from other lineages. The Omicron variants in the cluster are further formed into two groups with 100% bootstrap support, in which one merely contains a greater number of 21K (194) and the other one has 21L and 22C that are the most commonly circulating variants presently. The same viral lineage in the phylogenetic cluster was found in patients from two universities (i.e. Omicron, 21K), confirming the widespread transmission of the virus within university populations. The metadata includes samples that were specific to each of the two universities (purple), gender (orange), and year of sample collection (cyan), which are visualized as circles exterior to the branches in the tree (Figure 1; Table 1). This data reveals that i) the majority (62%, 335) of the samples studied were from University of Wisconsin-Milwaukee, ii) the majority of the samples (64%, 342) were collected in 2021, iii) while 81 of 537 samples have no gender/age data, a maximum (49%, 261) number of samples were collected from females, and iv) age of the patients ranged from 18-43 years with a median of 21 years.

**Table 1.** Characteristics of two university populations in Milwaukee, Wisconsin, United States, September 2020-April 2022.

| Characteristic |  | Marquette University | University of Wisconsin-Milwaukee | Total |
| --- | --- | --- | --- | --- |
| No. of samples sequenced |  | 202 | 335 | 537 |
| Gender |  |  |  |  |
|  | Female | 85 | 176 | 261 |
|  | Male | 62 | 133 | 195 |
|  | Unknown | 55 | 26 | 81 |
| Age* |  |  |  |  |
|  | Female | 18-35 (21) | 18-40 (21) | 18-40 (21) |
|  | Male | 18-35 (22) | 18-43 (22) | 18-43 (22) |

\*Value in the parenthesis is a median.

The comparison of the universities’ SARS-CoV-2 variants with the existing Global Initiative on Sharing All Influenza Data (GISAID) data revealed that the overall pattern of distributions of SARS-CoV-2 lineages tracked with national trends (Figure S3; Table S1) [21-23]. To discover the transmission ability of specific SARS-CoV-2 strains circulating in the university populations, we identified the existing genetically closely related genomes for Omicron BA.1/BA.2 and their descendent lineages, which were detected initially in each of two university samples (Table 2). This analysis along with phylogenetic clusters had identified that potential virus transmission occurred within, as well as between, the two university populations (i.e., BA.1.20, EPI_ISL_7744418 and BA.1, EPI_ISL_8477776 strains) (Table 2; Figure S4). Some strains of descendent lineages (i.e., BA.2.9, EPI_ISL_12431125; BA.1.1.18, EPI_ISL_7877072) in the university samples exhibited that they were detected in the university prior to the first widespread local community’s detection and vice versa (i.e., BA.1.15, EPI_ISL_8477784; BA.2, EPI_ISL_11301566) (Table 2), indicating that patterns of SARS-CoV-2 introduction and spread may differ considerably in the universities and communities. This analysis also reveals that some specific strains might have emerged (BA.1.20, EPI_ISL_7744418) from the university or disappeared (i.e., BA.2.3, EPI_ISL_12746651) without further transmission in the university and the local Milwaukee community. Altogether this result demonstrates that disease spreading in the university and community was possibly due to multiple independent introductions of different SARS-CoV-2 strains of variants over the periods, and as expected a few strains may have successfully established to spread within the university population, and also between universities and/or communities.

**Table 2.**
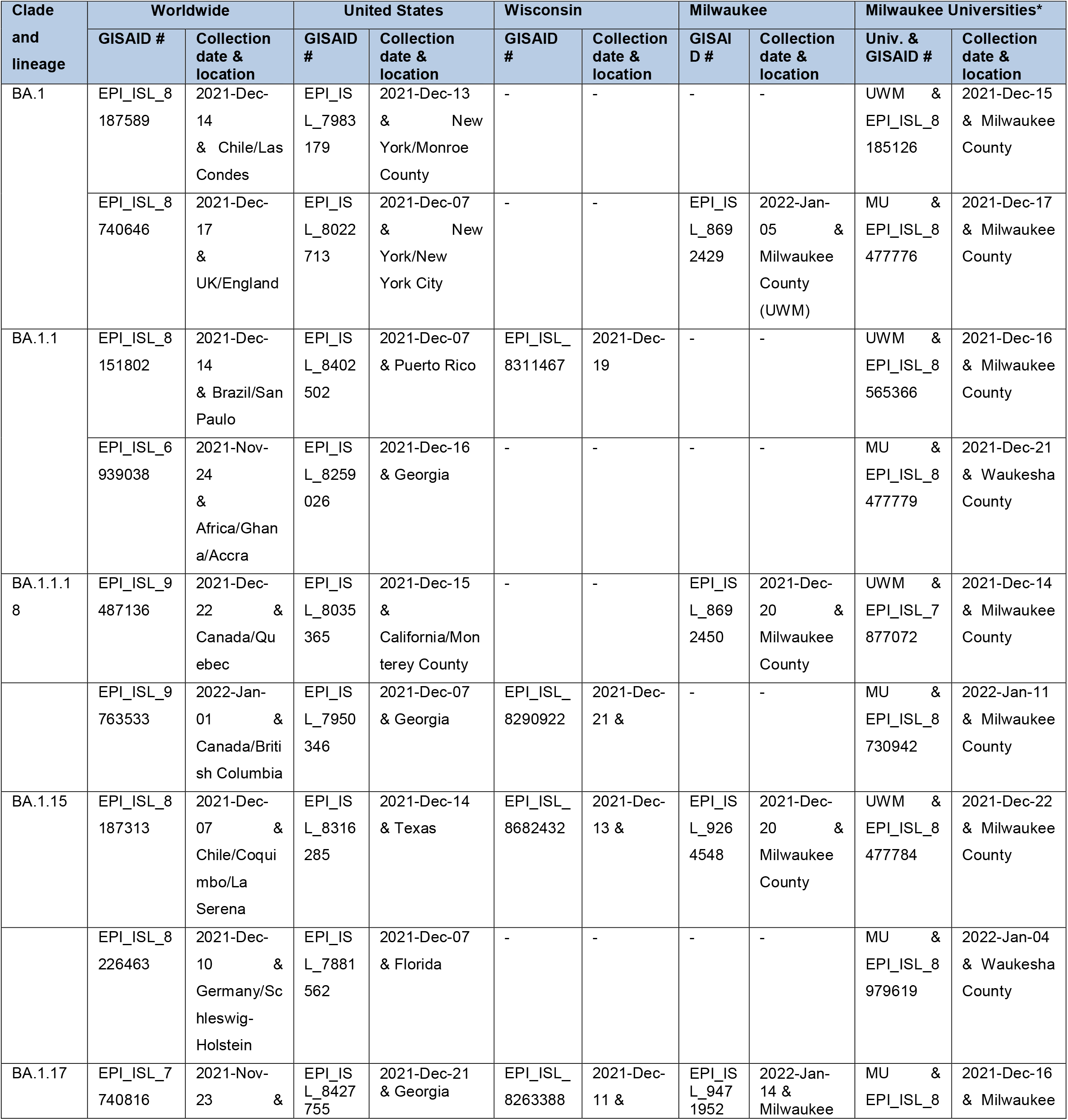
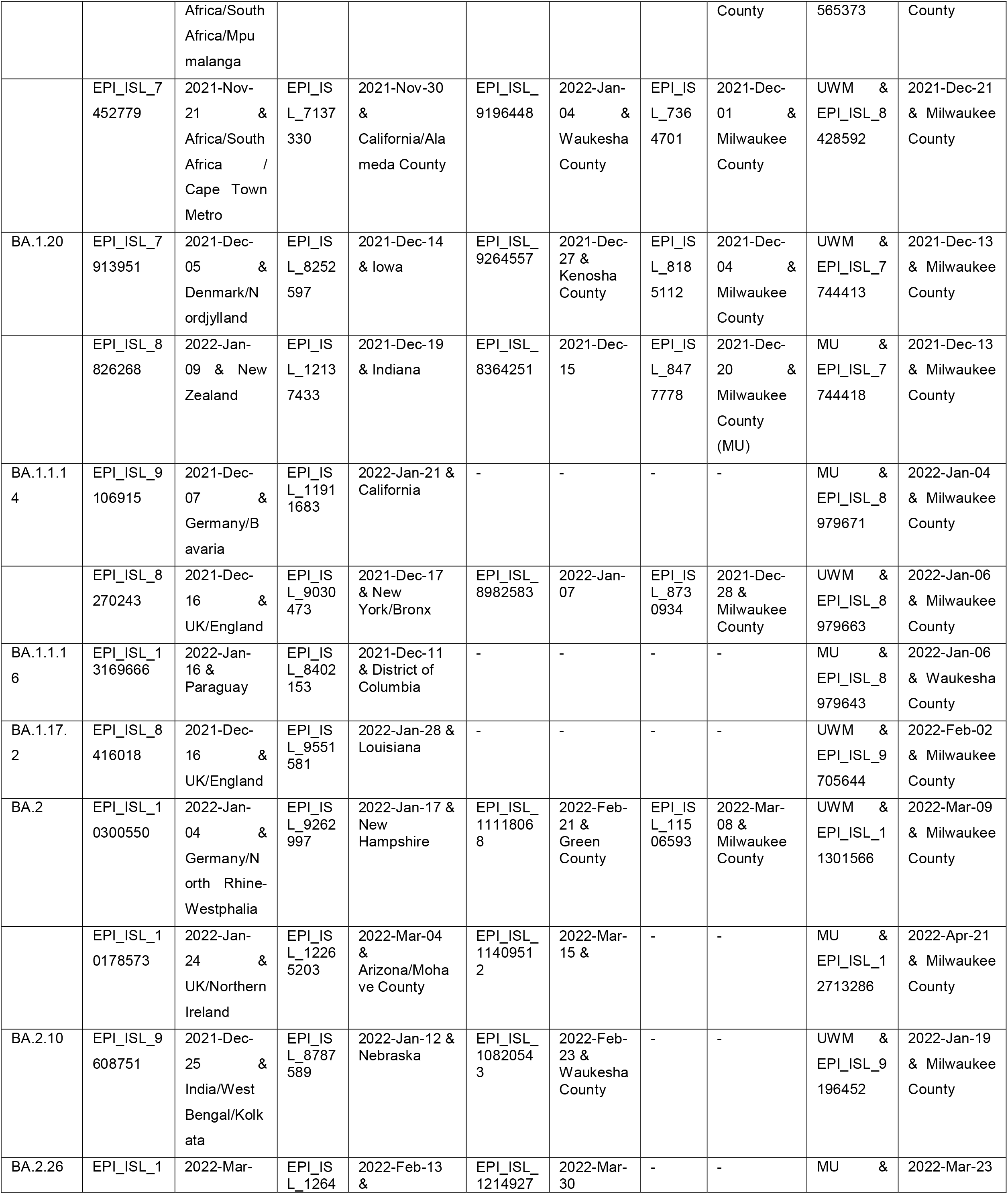
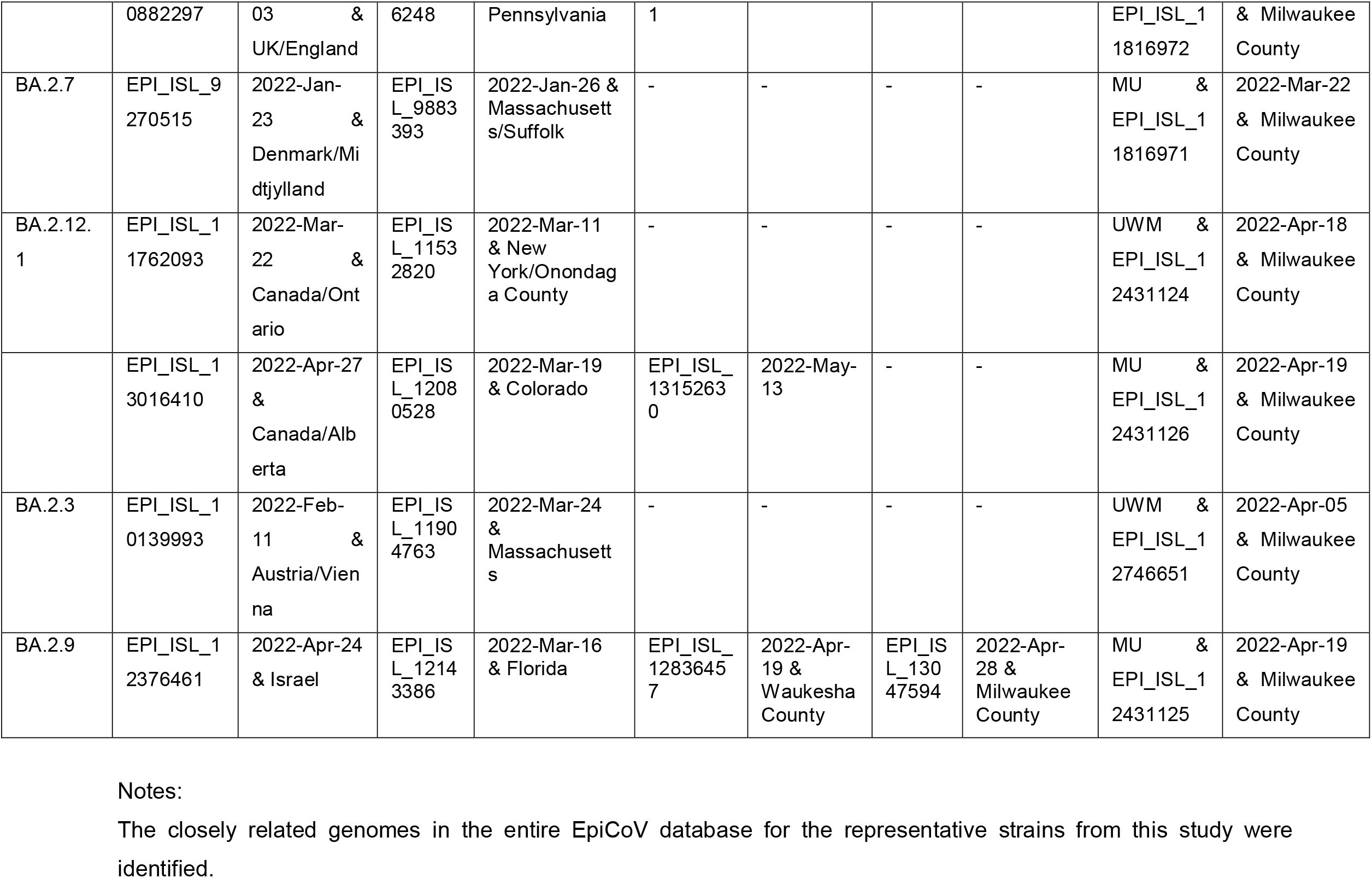
Details of closely related SARS-CoV-2 genomes of circulating Omicron strains identified among two university populations in the Milwaukee area. (public data accessed on June 21, 2022).

## Conclusions

Since June 3, 2022 the COVID-19 positive cases in the City of Milwaukee have increased to 14.1%, and it has moved into the extreme transmission category. Per CDC criteria, Milwaukee County moved into the high COVID-19 community level. Currently, 64.8% of City of Milwaukee adults 16 years and older are fully vaccinated, and 49.1% of fully vaccinated individuals have received a booster dose [24], apparently indicating that a significant portion of unvaccinated and/or partially vaccinated population in Milwaukee, along with unmasked gatherings and unrestricted traveling, are potentially influencing the rapid SARS-CoV-2 transmission in the community. Likewise, in-person learning, on-campus housing, off-campus university populations, and major sporting events created potential for rapid transmission of emerging variants (i.e. Omicron lineages) among the university’s populations. MHDL molecular surveillance efforts contributed in the identification of outbreak clusters on these campuses and have helped the universities to monitor and track suspected cases. While this study provides some insight into the patterns of SARS-CoV-2 introduction and spread in the university populations and the local community based on the available sequences, it only represents a portion of the viral transmission chains and is not a true estimation of the total circulating SARS-CoV-2 population. Further, expanding real-time sequencing, bioinformatics and data analysis capacity will aid local jurisdictions not only in identifying emerging SARS-CoV-2 variants like BA.4 and BA.5, but improve data-driven public health mitigation and policy efforts.

## Data Availability

All sequences data have been deposited with the GISAID. The accession number for all these sequences can be found on supplemental data file.

## Acknowledgments

Authors are grateful to the local university partners, AIDS Virus Research Laboratory (AVRL), Wisconsin State Lab of Hygiene and Wisconsin Department of Health Services staff for their support. Special thanks to Professors David O’Connor and Tom Friedrich at AVRL, UW-Madison and Dr. Heather Paradis at the Milwaukee Health Department Laboratory (MHDL) for their careful review of the manuscript. We acknowledge the critical role of the Milwaukee Health Department (MHD) communicable disease and preparedness team in coordinating respiratory specimen collection, case investigations, and contact tracing. Thanks to the Milwaukee Health Department Laboratory (MHDL) staff who were involved in order entry, extraction of nucleic acids, SARS-CoV-2 PCR setup, and result reporting. Special thanks to Addie Skillman, Samantha Scott, Jennifer Lentz, and Nandhakumar Balakrishnan for their technical assistance in the laboratory, Noah Leigh for LIS support and Kristin Schieble for the laboratory operations coordination. We appreciate all of the support from the MHD administration and leadership team during this study. We gratefully acknowledge the authors, the originating and submitting Laboratories for their sequence and metadata shared through GISAID and GenBank. The manuscript contents are those of the authors and do not necessarily represent the official views of, nor an endorsement, by the City of Milwaukee Health Department.

## Author Contributions

AR, MK and SB conceptualized the study. MK, AB, KA, JW performed experiments, and genome sequencing. AR performed bioinformatics analysis. AR, MK, AB, KA, JW and SB involved in data interpretation. AR wrote the manuscript with input from all authors.

---

**Figure S1.**
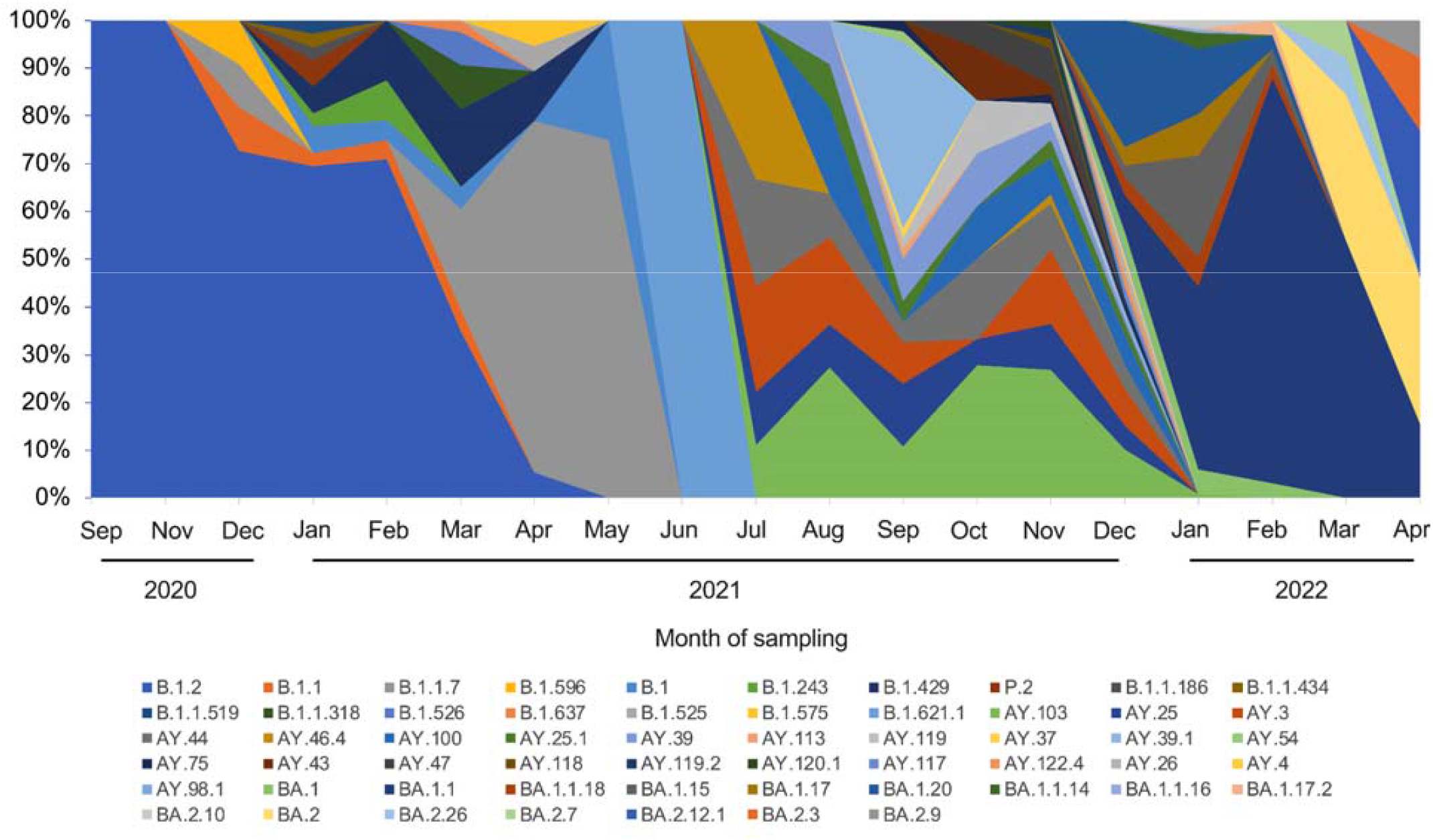
Chronologic distribution of SARS-CoV-2 genomic variants for 20 months in the Milwaukee university’s populations. The lineage and sub-lineage of 537 genome sequences of SARS-CoV-2 were identified in Nextclade and classified based on the sampling date from September 2020-April 2022. The majority (>50%) of the SARS-CoV-2 viruses belonged to 6 sub-lineages: BA.1.1 (88, 16.39%), B.1.2 (74, 13.78%), BA.1.20 (38, 7.08%), AY.103 (37, 6.89%), BA.1.15 (28, 5.21%), and B.1.1.7 (27, 5.03%).

**Figure S2.**
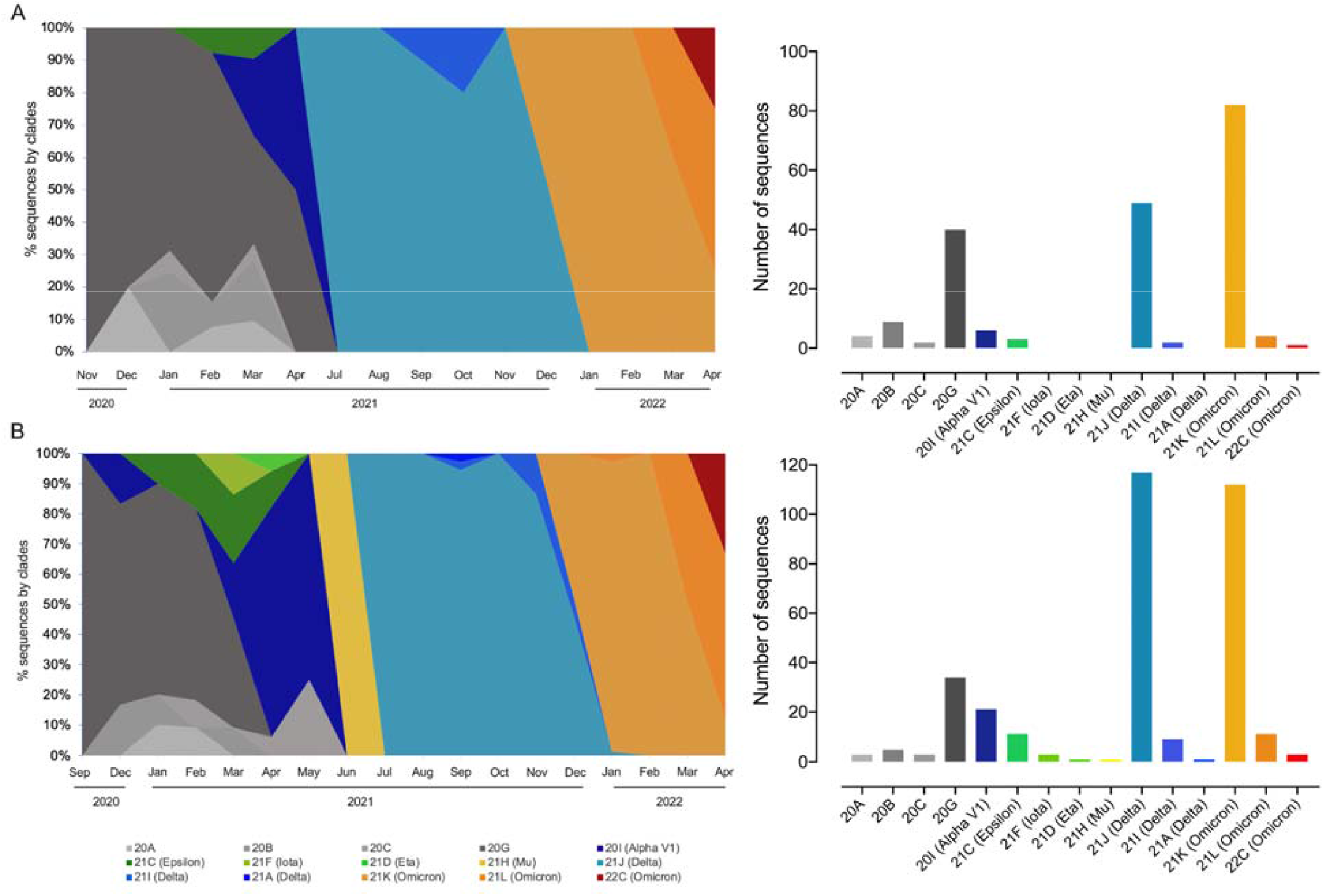
Independent assessment of the chronologic distributions of SARS-CoV-2 sequences from September 2020-April 2022 in two universities: A) Marquette University and B) University of Wisconsin-Milwaukee. Cumulative number of sequences segregated based on the Nextclade classification are provided in the right panels.

**Figure S3.**
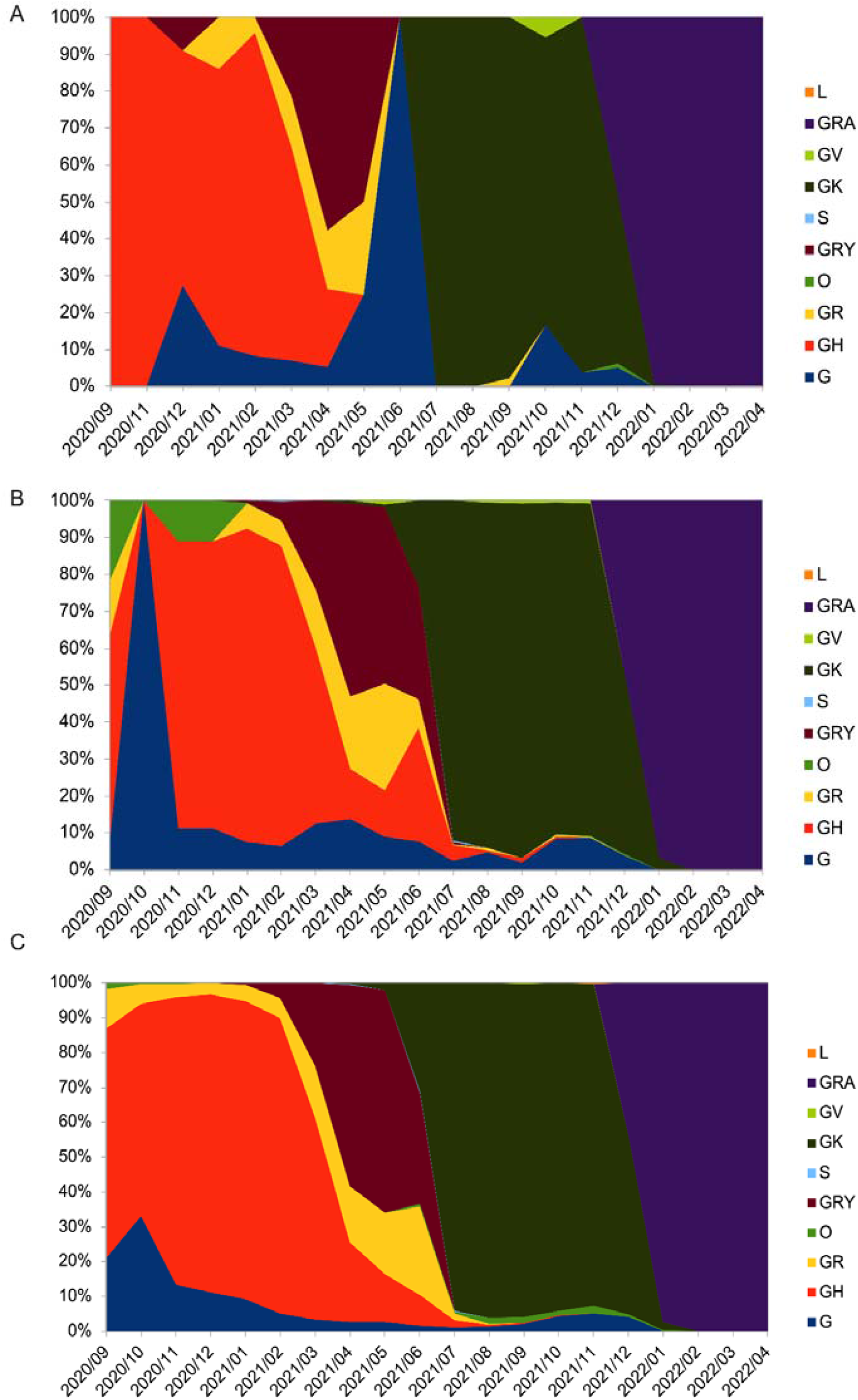
Comparison of chronologic distribution of SARS-CoV-2 genomic variants (as per GISAID clade classification) sampled from September 2020-April 2022. A) 537 sequences from Milwaukee universities populations (this study), B) 4,375 sequences from Milwaukee County populations (GISAID), and C) 57,365 sequences from Wisconsin State populations (GISAID).

**Figure S4.**
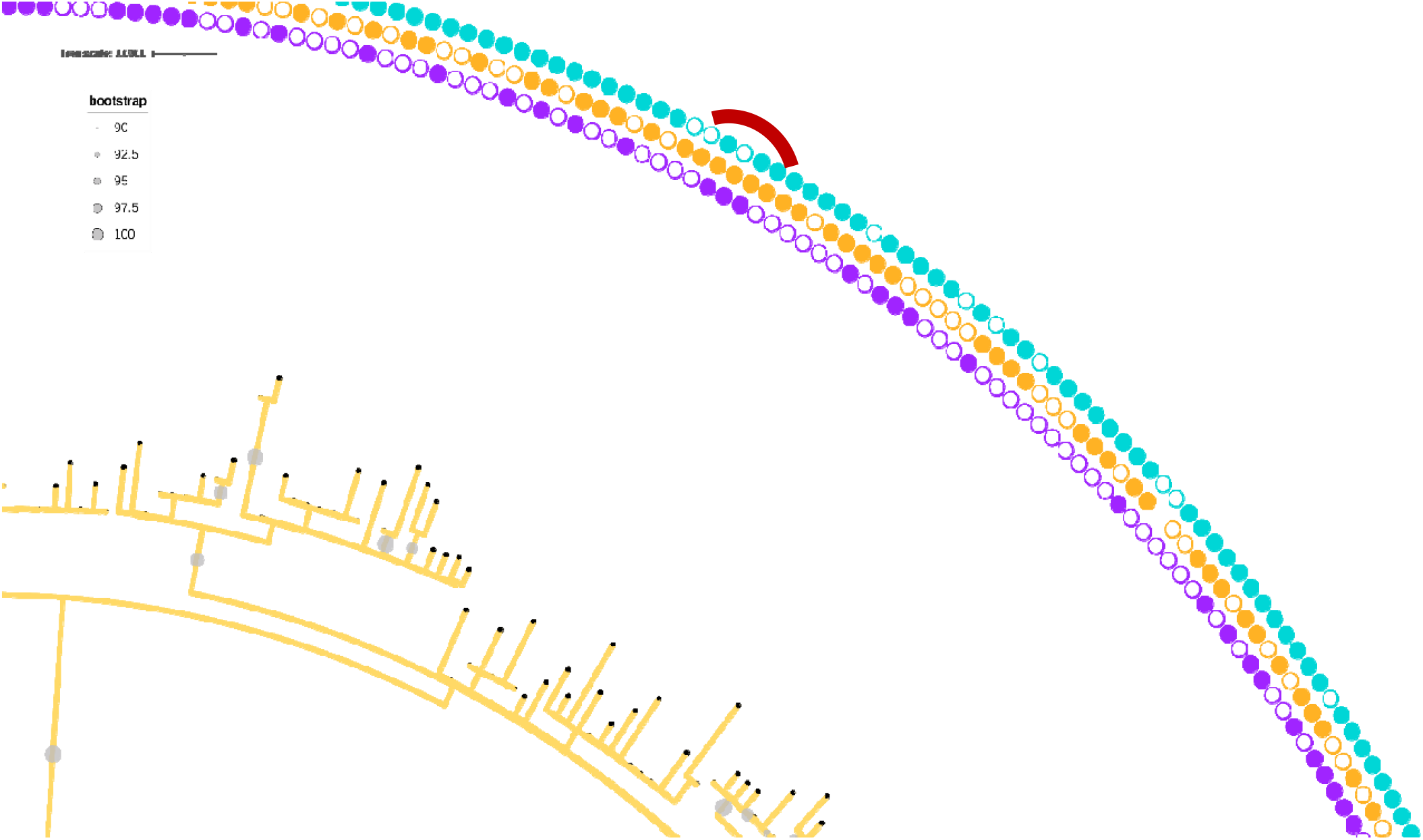
Representative phylogenetic cluster (red curve) highlighting the transmission within and between the universities. The metadata are visualized as circles exterior to the branches in the tree. From the inner to the outer concentric circle of the branches: Circle 1–3 are indicated samples that are specific to each of two universities, gender and year of sample collection, respectively. In circle 1 (purple), the filled and outlined circle symbols exemplify that the clinical samples used for sequencing originated from Marquette University (MU) and University of Wisconsin-Milwaukee (UWM), respectively. In circle 2 (orange), gender female (filled circle symbol)) and male (outlined) shown, while, the absence of symbol indicated that no gender data available for the given samples. While the filled and outlined circle symbols in the circle 3 (cyan) indicated the years 2022 and 2021, respectively, the absence of symbol indicated 2020, in which the samples were collected from the patients.

**Table S1.**
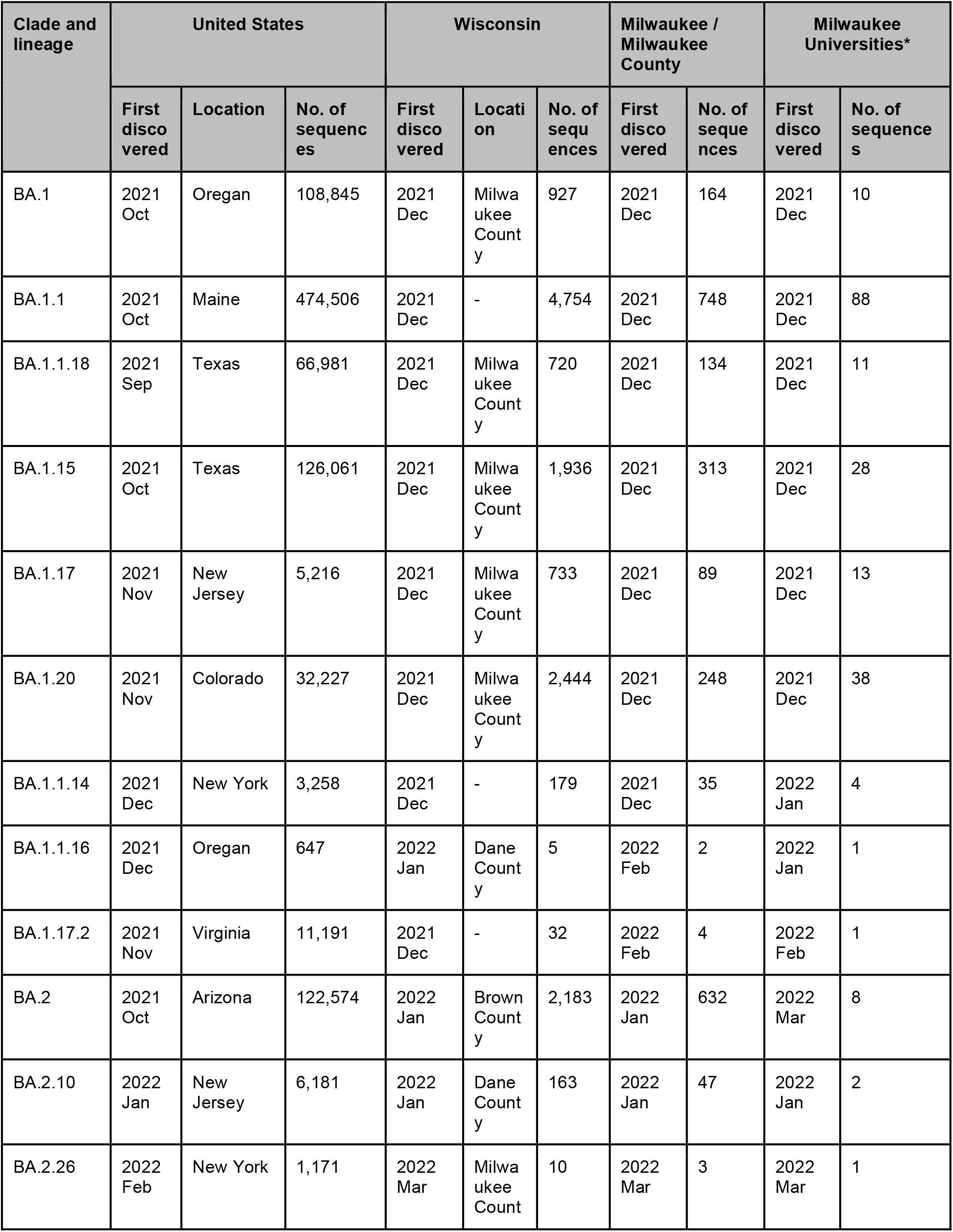
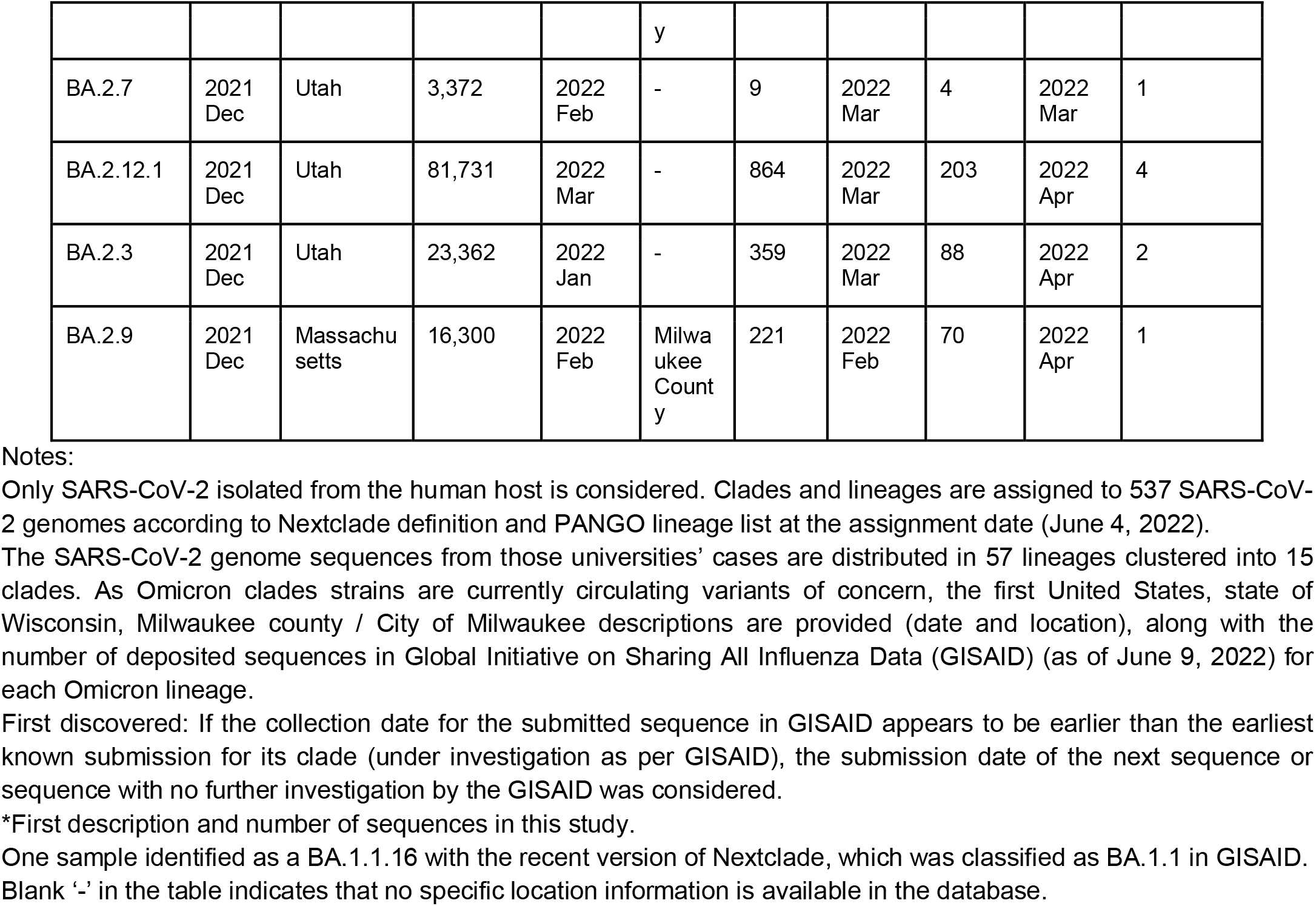
Circulating Omicron clades and descendent lineages identified among two university populations in Milwaukee County/ City of Milwaukee, Wisconsin, United States, September 2020-April 2022.

