## Supplemental Data for "Genomic Surveillance Identifies SARS-CoV-2 Transmission Patterns in Local University Populations, Wisconsin, USA, 2020–2022"

**Supplemental data: 537 SARS-CoV-2 details.**

| Virus_name | EPI_ISL_ID |
|---|---|
| hCoV-19/USA/WI-MHDL-8122101/2021 | EPI_ISL_3836426 |
| hCoV-19/USA/WI-MHDL-8312104/2021 | EPI_ISL_4552705 |
| hCoV-19/USA/WI-MHDL-202110_0047/2021 | EPI_ISL_5850493 |
| hCoV-19/USA/WI-MHDL-202110_0037/2021 | EPI_ISL_5850485 |
| hCoV-19/USA/WI-MHDL-2021110436/2021 | EPI_ISL_6861948 |
| hCoV-19/USA/WI-MHDL-2021110447/2021 | EPI_ISL_6861957 |
| hCoV-19/USA/WI-MHDL-2021110453/2021 | EPI_ISL_6864926 |
| hCoV-19/USA/WI-MHDL-2021110364/2021 | EPI_ISL_6945701 |
| hCoV-19/USA/WI-MHDL-2021120144/2021 | EPI_ISL_7809497 |
| hCoV-19/USA/WI-MHDL-2021120140/2021 | EPI_ISL_7809493 |
| hCoV-19/USA/WI-MHDL-2021120151/2021 | EPI_ISL_7809504 |
| hCoV-19/USA/WI-MHDL-2021120311/2021 | EPI_ISL_7828371 |
| hCoV-19/USA/WI-MHDL-2021120503/2021 | EPI_ISL_8185129 |
| hCoV-19/USA/WI-MHDL-7302102/2021 | EPI_ISL_3535698 |
| hCoV-19/USA/WI-MHDL-991327860/2021 | EPI_ISL_3836428 |
| hCoV-19/USA/WI-MHDL-991334437/2021 | EPI_ISL_3893751 |
| hCoV-19/USA/WI-MHDL-006206684/2021 | EPI_ISL_4552703 |
| hCoV-19/USA/WI-MHDL-U9100019/2021 | EPI_ISL_4552723 |
| hCoV-19/USA/WI-MHDL-202109_0099/2021 | EPI_ISL_5053445 |
| hCoV-19/USA/WI-MHDL-202109_0107/2021 | EPI_ISL_5053453 |
| hCoV-19/USA/WI-MHDL-202110_0010/2021 | EPI_ISL_5053480 |
| hCoV-19/USA/WI-MHDL-202110_0001/2021 | EPI_ISL_5053471 |
| hCoV-19/USA/WI-MHDL-202110_0048/2021 | EPI_ISL_5850494 |
| hCoV-19/USA/WI-MHDL-202110_0054/2021 | EPI_ISL_5850500 |
| hCoV-19/USA/WI-MHDL-202110_0038/2021 | EPI_ISL_5850486 |
| hCoV-19/USA/WI-MHDL-2021110189/2021 | EPI_ISL_6864869 |
| hCoV-19/USA/WI-MHDL-2021110211/2021 | EPI_ISL_6594792 |
| hCoV-19/USA/WI-MHDL-2021110223/2021 | EPI_ISL_6594800 |
| hCoV-19/USA/WI-MHDL-2021110195/2021 | EPI_ISL_6864872 |
| hCoV-19/USA/WI-MHDL-2021110194/2021 | EPI_ISL_6864871 |
| hCoV-19/USA/WI-MHDL-2021110441/2021 | EPI_ISL_6861953 |
| hCoV-19/USA/WI-MHDL-2021110442/2021 | EPI_ISL_6861954 |
| hCoV-19/USA/WI-MHDL-2021110472/2021 | EPI_ISL_6861980 |
| hCoV-19/USA/WI-MHDL-2021110456/2021 | EPI_ISL_6861965 |
| hCoV-19/USA/WI-MHDL-2021110480/2021 | EPI_ISL_6861988 |
| hCoV-19/USA/WI-MHDL-2021120020/2021 | EPI_ISL_7153332 |
| hCoV-19/USA/WI-MHDL-2021120045/2021 | EPI_ISL_7281475 |
| hCoV-19/USA/WI-MHDL-2021120046/2021 | EPI_ISL_7281483 |
| hCoV-19/USA/WI-MHDL-2021120050/2021 | EPI_ISL_7281516 |
| hCoV-19/USA/WI-MHDL-2021120049/2021 | EPI_ISL_7281507 |

| | |
|---|---|
| hCoV-19/USA/WI-MHDL-2021120047/2021 | EPI_ISL_7281492 |
| hCoV-19/USA/WI-MHDL-2021120143/2021 | EPI_ISL_7809496 |
| hCoV-19/USA/WI-MHDL-2021120196/2021 | EPI_ISL_7478637 |
| hCoV-19/USA/WI-MHDL-2021120369/2021 | EPI_ISL_7744419 |
| hCoV-19/USA/WI-MHDL-2021120366/2021 | EPI_ISL_7744416 |
| hCoV-19/USA/WI-MHDL-2021120384/2021 | EPI_ISL_7877070 |
| hCoV-19/USA/WI-MHDL-2021120501/2021 | EPI_ISL_8185127 |
| hCoV-19/USA/WI-MHDL-2022010023/2021 | EPI_ISL_8388844 |
| hCoV-19/USA/WI-MHDL-2022010020/2021 | EPI_ISL_8388841 |
| hCoV-19/USA/WI-MHDL-2022010258/2022 | EPI_ISL_8979610 |
| hCoV-19/USA/WI-MHDL-U9100012/2021 | EPI_ISL_4552727 |
| hCoV-19/USA/WI-MHDL-2021120202/2021 | EPI_ISL_7660960 |
| hCoV-19/USA/WI-MHDL-2021120302/2021 | EPI_ISL_7828365 |
| hCoV-19/USA/WI-MHDL-2021110438/2021 | EPI_ISL_6861950 |
| hCoV-19/USA/WI-MHDL-202109_0104/2021 | EPI_ISL_5053450 |
| hCoV-19/USA/WI-MHDL-202110_0060/2021 | EPI_ISL_5850505 |
| hCoV-19/USA/WI-MHDL-202110_0065/2021 | EPI_ISL_5850510 |
| hCoV-19/USA/WI-MHDL-2021110451/2021 | EPI_ISL_6861961 |
| hCoV-19/USA/WI-MHDL-2021120048/2021 | EPI_ISL_7281499 |
| hCoV-19/USA/WI-MHDL-2021110336/2021 | EPI_ISL_6571154 |
| hCoV-19/USA/WI-MHDL-2022010173/2021 | EPI_ISL_8565368 |
| hCoV-19/USA/WI-MHDL-2021110448/2021 | EPI_ISL_6861958 |
| hCoV-19/USA/WI-MHDL-2021120203/2021 | EPI_ISL_7660962 |
| hCoV-19/USA/WI-MHDL-2021120351/2021 | EPI_ISL_7698329 |
| hCoV-19/USA/WI-MHDL-006170030/2021 | EPI_ISL_3535708 |
| hCoV-19/USA/WI-MHDL-991221498/2021 | EPI_ISL_4552706 |
| hCoV-19/USA/WI-MHDL-9012119/2021 | EPI_ISL_4552707 |
| hCoV-19/USA/WI-MHDL-006198912/2021 | EPI_ISL_4552709 |
| hCoV-19/USA/WI-MHDL-006148902/2021 | EPI_ISL_4552724 |
| hCoV-19/USA/WI-MHDL-9032107/2021 | EPI_ISL_4552718 |
| hCoV-19/USA/WI-MHDL-006153142/2021 | EPI_ISL_4552710 |
| hCoV-19/USA/WI-MHDL-202110_0028/2021 | EPI_ISL_5419466 |
| hCoV-19/USA/WI-MHDL-202110_0036/2021 | EPI_ISL_5850484 |
| hCoV-19/USA/WI-MHDL-2021110471/2021 | EPI_ISL_6861979 |
| hCoV-19/USA/WI-MHDL-2021110366/2021 | EPI_ISL_6945703 |
| hCoV-19/USA/WI-MHDL-2021110368/2021 | EPI_ISL_6945705 |
| hCoV-19/USA/WI-MHDL-2021110369/2021 | EPI_ISL_6945706 |
| hCoV-19/USA/WI-MHDL-2021120029/2021 | EPI_ISL_7153338 |
| hCoV-19/USA/WI-MHDL-2021120142/2021 | EPI_ISL_7809495 |
| hCoV-19/USA/WI-MHDL-2021120145/2021 | EPI_ISL_7809498 |
| hCoV-19/USA/WI-MHDL-2021120163/2021 | EPI_ISL_7364692 |
| hCoV-19/USA/WI-MHDL-2021120161/2021 | EPI_ISL_7364690 |
| hCoV-19/USA/WI-MHDL-8172111/2021 | EPI_ISL_3836427 |

| | |
|---|---|
| hCoV-19/USA/WI-MHDL-202110110093/2021 | EPI_ISL_5330265 |
| hCoV-19/USA/WI-MHDL-202110110094/2021 | EPI_ISL_5330264 |
| hCoV-19/USA/WI-MHDL-2021110335/2021 | EPI_ISL_6571153 |
| hCoV-19/USA/WI-MHDL-2021110365/2021 | EPI_ISL_6945702 |
| hCoV-19/USA/WI-MHDL-2021120148/2021 | EPI_ISL_7809501 |
| hCoV-19/USA/WI-MHDL-2021120160/2021 | EPI_ISL_7364689 |
| hCoV-19/USA/WI-MHDL-2021120162/2021 | EPI_ISL_7364691 |
| hCoV-19/USA/WI-MHDL-2021120195/2021 | EPI_ISL_7478636 |
| hCoV-19/USA/WI-MHDL-006147146/2021 | EPI_ISL_3535704 |
| hCoV-19/USA/WI-MHDL-006147634/2021 | EPI_ISL_3535707 |
| hCoV-19/USA/WI-MHDL-8022107/2021 | EPI_ISL_3535700 |
| hCoV-19/USA/WI-MHDL-8042117/2021 | EPI_ISL_3535701 |
| hCoV-19/USA/WI-MHDL-005804741/2021 | EPI_ISL_4552704 |
| hCoV-19/USA/WI-MHDL-991392191/2021 | EPI_ISL_4552720 |
| hCoV-19/USA/WI-MHDL-202109_0098/2021 | EPI_ISL_5053444 |
| hCoV-19/USA/WI-MHDL-202110_0002/2021 | EPI_ISL_5053472 |
| hCoV-19/USA/WI-MHDL-2021110435/2021 | EPI_ISL_6861947 |
| hCoV-19/USA/WI-MHDL-2021110219/2021 | EPI_ISL_6594796 |
| hCoV-19/USA/WI-MHDL-2021110439/2021 | EPI_ISL_6861951 |
| hCoV-19/USA/WI-MHDL-2021110440/2021 | EPI_ISL_6861952 |
| hCoV-19/USA/WI-MHDL-2021110449/2021 | EPI_ISL_6861959 |
| hCoV-19/USA/WI-MHDL-2021110445/2021 | EPI_ISL_6861955 |
| hCoV-19/USA/WI-MHDL-2021110446/2021 | EPI_ISL_6861956 |
| hCoV-19/USA/WI-MHDL-2021110481/2021 | EPI_ISL_6861989 |
| hCoV-19/USA/WI-MHDL-2021120149/2021 | EPI_ISL_7809502 |
| hCoV-19/USA/WI-MHDL-2021120197/2021 | EPI_ISL_7478638 |
| hCoV-19/USA/WI-MHDL-2021120383/2021 | EPI_ISL_7744430 |
| hCoV-19/USA/WI-MHDL-2021120367/2021 | EPI_ISL_7744417 |
| hCoV-19/USA/WI-MHDL-2021120365/2021 | EPI_ISL_7744415 |
| hCoV-19/USA/WI-MHDL-2021120391/2021 | EPI_ISL_7877061 |
| hCoV-19/USA/WI-MHDL-202110_0009/2021 | EPI_ISL_5053479 |
| hCoV-19/USA/WI-MHDL-991271039/2021 | EPI_ISL_4552719 |
| hCoV-19/USA/WI-MHDL-006230272/2021 | EPI_ISL_4552726 |
| hCoV-19/USA/WI-MHDL-202110_0008/2021 | EPI_ISL_5053478 |
| hCoV-19/USA/WI-MHDL-202110_0007/2021 | EPI_ISL_5053477 |
| hCoV-19/USA/WI-MHDL-202110_0004/2021 | EPI_ISL_5053474 |
| hCoV-19/USA/WI-MHDL-202110_0049/2021 | EPI_ISL_5850495 |
| hCoV-19/USA/WI-MHDL-202110_0059/2021 | EPI_ISL_5850504 |
| hCoV-19/USA/WI-MHDL-2021110454/2021 | EPI_ISL_6861963 |
| hCoV-19/USA/WI-MHDL-2021110482/2021 | EPI_ISL_6861990 |
| hCoV-19/USA/WI-MHDL-2021120446/2021 | EPI_ISL_7979270 |
| hCoV-19/USA/WI-MHDL-991412080/2021 | EPI_ISL_4552711 |
| hCoV-19/USA/WI-MHDL-991406185/2021 | EPI_ISL_4552712 |

| | |
|---|---|
| hCoV-19/USA/WI-MHDL-991398784/2021 | EPI_ISL_4552717 |
| hCoV-19/USA/WI-MHDL-9082140/2021 | EPI_ISL_4552713 |
| hCoV-19/USA/WI-MHDL-9082126/2021 | EPI_ISL_4552716 |
| hCoV-19/USA/WI-MHDL-U9090002/2021 | EPI_ISL_4552728 |
| hCoV-19/USA/WI-MHDL-9082104/2021 | EPI_ISL_4552714 |
| hCoV-19/USA/WI-MHDL-9092128/2021 | EPI_ISL_4552702 |
| hCoV-19/USA/WI-MHDL-U9100006/2021 | EPI_ISL_4552722 |
| hCoV-19/USA/WI-MHDL-U9100041/2021 | EPI_ISL_4552725 |
| hCoV-19/USA/WI-MHDL-U9100022/2021 | EPI_ISL_4552729 |
| hCoV-19/USA/WI-MHDL-202109_0102/2021 | EPI_ISL_5053448 |
| hCoV-19/USA/WI-MHDL-U9130042/2021 | EPI_ISL_4552730 |
| hCoV-19/USA/WI-MHDL-202109_0097/2021 | EPI_ISL_5053443 |
| hCoV-19/USA/WI-MHDL-U9140190/2021 | EPI_ISL_4552721 |
| hCoV-19/USA/WI-MHDL-202109_0108/2021 | EPI_ISL_5053454 |
| hCoV-19/USA/WI-MHDL-202110_0011/2021 | EPI_ISL_5053481 |
| hCoV-19/USA/WI-MHDL-202110_0005/2021 | EPI_ISL_5053475 |
| hCoV-19/USA/WI-MHDL-2021120370/2021 | EPI_ISL_7744420 |
| hCoV-19/USA/WI-MHDL-2022010171/2021 | EPI_ISL_8565367 |
| hCoV-19/USA/WI-MHDL-202110_0052/2021 | EPI_ISL_5850498 |
| hCoV-19/USA/WI-MHDL-202110_0053/2021 | EPI_ISL_5850499 |
| hCoV-19/USA/WI-MHDL-2021110367/2021 | EPI_ISL_6945704 |
| hCoV-19/USA/WI-MHDL-7262106/2021 | EPI_ISL_3535697 |
| hCoV-19/USA/WI-MHDL-006136991/2021 | EPI_ISL_3535706 |
| hCoV-19/USA/WI-MHDL-5092103/2021 | EPI_ISL_3535702 |
| hCoV-19/USA/WI-MHDL-006155116/2021 | EPI_ISL_4552701 |
| hCoV-19/USA/WI-MHDL-9072113/2021 | EPI_ISL_4552708 |
| hCoV-19/USA/WI-MHDL-202110_0057/2021 | EPI_ISL_5850502 |
| hCoV-19/USA/WI-MHDL-2021110450/2021 | EPI_ISL_6861960 |
| hCoV-19/USA/WI-MHDL-202110_0058/2021 | EPI_ISL_5850503 |
| hCoV-19/USA/WI-MHDL-2021110220/2021 | EPI_ISL_6594797 |
| hCoV-19/USA/WI-MHDL-2021110485/2021 | EPI_ISL_6861993 |
| hCoV-19/USA/WI-MHDL-2021110483/2021 | EPI_ISL_6861991 |
| hCoV-19/USA/WI-MHDL-2021120146/2021 | EPI_ISL_7809499 |
| hCoV-19/USA/WI-MHDL-2021120147/2021 | EPI_ISL_7809500 |
| hCoV-19/USA/WI-MHDL-2021120141/2021 | EPI_ISL_7809494 |
| hCoV-19/USA/WI-MHDL-2021120150/2021 | EPI_ISL_7809503 |
| hCoV-19/USA/WI-MHDL-2021120382/2021 | EPI_ISL_7744429 |
| hCoV-19/USA/WI-MHDL-2022010022/2021 | EPI_ISL_8388842 |
| hCoV-19/USA/WI-MHDL-005769138/2021 | EPI_ISL_3535703 |
| hCoV-19/USA/WI-MHDL-006210158/2021 | EPI_ISL_3535705 |
| hCoV-19/USA/WI-MHDL-7302103/2021 | EPI_ISL_3535699 |
| hCoV-19/USA/WI-MHDL-2021110196/2021 | EPI_ISL_6864873 |
| hCoV-19/USA/WI-MHDL-202110_0039/2021 | EPI_ISL_5850487 |

| | |
|---|---|
| hCoV-19/USA/WI-MHDL-2021110224/2021 | EPI_ISL_6594801 |
| hCoV-19/USA/WI-MHDL-2021110437/2021 | EPI_ISL_6861949 |
| hCoV-19/USA/WI-MHDL-2021110455/2021 | EPI_ISL_6861964 |
| hCoV-19/USA/WI-MHDL-2021120021/2021 | EPI_ISL_7153335 |
| hCoV-19/USA/WI-MHDL-202110_0006/2021 | EPI_ISL_5053476 |
| hCoV-19/USA/WI-MHDL-202109_0103/2021 | EPI_ISL_5053449 |
| hCoV-19/USA/WI-MHDL-2021110363/2021 | EPI_ISL_6945700 |
| hCoV-19/USA/WI-MHDL-2021120364/2021 | EPI_ISL_7744414 |
| hCoV-19/USA/WI-MHDL-2021120209/2021 | EPI_ISL_7660966 |
| hCoV-19/USA/WI-MHDL-0672/2021 | EPI_ISL_1494754 |
| hCoV-19/USA/WI-MHDL-MU0271/2021 | EPI_ISL_1493388 |
| hCoV-19/USA/WI-MHDL-U2170021/2021 | EPI_ISL_1262589 |
| hCoV-19/USA/WI-MHDL-MU0309/2021 | EPI_ISL_1652155 |
| hCoV-19/USA/WI-MHDL-MU0313/2021 | EPI_ISL_1652158 |
| hCoV-19/USA/WI-MHDL-5042117/2021 | EPI_ISL_2232212 |
| hCoV-19/USA/WI-MHDL-0634/2020 | EPI_ISL_1592250 |
| hCoV-19/USA/WI-MHDL-0664/2021 | EPI_ISL_1652182 |
| hCoV-19/USA/WI-MHDL-MU0278/2021 | EPI_ISL_1262594 |
| hCoV-19/USA/WI-MHDL-0702/2021 | EPI_ISL_1592240 |
| hCoV-19/USA/WI-MHDL-0704/2021 | EPI_ISL_1592242 |
| hCoV-19/USA/WI-MHDL-MU0265/2021 | EPI_ISL_1494749 |
| hCoV-19/USA/WI-MHDL-MU0325/2021 | EPI_ISL_1608144 |
| hCoV-19/USA/WI-MHDL-MU0321/2021 | EPI_ISL_1608140 |
| hCoV-19/USA/WI-MHDL-MU0322/2021 | EPI_ISL_1608141 |
| hCoV-19/USA/WI-MHDL-MU0326/2021 | EPI_ISL_1608145 |
| hCoV-19/USA/WI-MPHL-0643/2021 | EPI_ISL_940734 |
| hCoV-19/USA/WI-MHDL-MU0268/2021 | EPI_ISL_1493385 |
| hCoV-19/USA/WI-MHDL-0641/2020 | EPI_ISL_1652173 |
| hCoV-19/USA/WI-MHDL-MU0311/2021 | EPI_ISL_1652157 |
| hCoV-19/USA/WI-MHDL-0687/2021 | EPI_ISL_1652160 |
| hCoV-19/USA/WI-MHDL-0689/2021 | EPI_ISL_1652162 |
| hCoV-19/USA/WI-MHDL-MU0318/2021 | EPI_ISL_1592244 |
| hCoV-19/USA/WI-MHDL-MU0320/2021 | EPI_ISL_1592246 |
| hCoV-19/USA/WI-MHDL-0703/2021 | EPI_ISL_1592241 |
| hCoV-19/USA/WI-MHDL-MU0323/2021 | EPI_ISL_1608142 |
| hCoV-19/USA/WI-MHDL-MU0324/2021 | EPI_ISL_1608143 |
| hCoV-19/USA/WI-MHDL-0708/2021 | EPI_ISL_1608147 |
| hCoV-19/USA/WI-MHDL-0712/2021 | EPI_ISL_1608150 |
| hCoV-19/USA/WI-MHDL-0714/2021 | EPI_ISL_1608152 |
| hCoV-19/USA/WI-MHDL-0715/2021 | EPI_ISL_1608153 |
| hCoV-19/USA/WI-MHDL-0716/2021 | EPI_ISL_1608154 |
| hCoV-19/USA/WI-MHDL-0718/2021 | EPI_ISL_1608156 |
| hCoV-19/USA/WI-MHDL-4162115/2021 | EPI_ISL_2179061 |

| | |
|---|---|
| hCoV-19/USA/WI-MHDL-4192101/2021 | EPI_ISL_2179064 |
| hCoV-19/USA/WI-MHDL-4192115/2021 | EPI_ISL_2179065 |
| hCoV-19/USA/WI-MHDL-4192109/2021 | EPI_ISL_2179062 |
| hCoV-19/USA/WI-MHDL-991311570/2021 | EPI_ISL_2179067 |
| hCoV-19/USA/WI-MHDL-4192112/2021 | EPI_ISL_2179066 |
| hCoV-19/USA/WI-MHDL-4192117/2021 | EPI_ISL_2179063 |
| hCoV-19/USA/WI-MHDL-006127339/2021 | EPI_ISL_2232210 |
| hCoV-19/USA/WI-MHDL-4262118/2021 | EPI_ISL_2230795 |
| hCoV-19/USA/WI-MHDL-5052107/2021 | EPI_ISL_2232213 |
| hCoV-19/USA/WI-MHDL-5062101/2021 | EPI_ISL_2233670 |
| hCoV-19/USA/WI-MHDL-U5260033/2021 | EPI_ISL_2876280 |
| hCoV-19/USA/WI-MPHL-0073/2020 | EPI_ISL_931474 |
| hCoV-19/USA/WI-MHDL-006194652/2020 | EPI_ISL_2179072 |
| hCoV-19/USA/WI-MHDL-006129641/2020 | EPI_ISL_2179073 |
| hCoV-19/USA/WI-MHDL-006133313/2020 | EPI_ISL_2230798 |
| hCoV-19/USA/WI-MHDL-006149715/2020 | EPI_ISL_2179075 |
| hCoV-19/USA/WI-MHDL-006188522/2020 | EPI_ISL_2179076 |
| hCoV-19/USA/WI-MHDL-006155587/2020 | EPI_ISL_2179077 |
| hCoV-19/USA/WI-MHDL-006148097/2020 | EPI_ISL_2230799 |
| hCoV-19/USA/WI-MHDL-MU0251/2020 | EPI_ISL_1652166 |
| hCoV-19/USA/WI-MHDL-MU0252/2020 | EPI_ISL_1652167 |
| hCoV-19/USA/WI-MHDL-MU0253/2020 | EPI_ISL_1652168 |
| hCoV-19/USA/WI-MHDL-MU0254/2020 | EPI_ISL_1652169 |
| hCoV-19/USA/WI-MHDL-0636/2020 | EPI_ISL_1592251 |
| hCoV-19/USA/WI-MHDL-0637/2020 | EPI_ISL_1592252 |
| hCoV-19/USA/WI-MHDL-0638/2020 | EPI_ISL_1652171 |
| hCoV-19/USA/WI-MHDL-0639/2020 | EPI_ISL_1652172 |
| hCoV-19/USA/WI-MPHL-0642/2021 | EPI_ISL_940733 |
| hCoV-19/USA/WI-MHDL-0659/2021 | EPI_ISL_1652179 |
| hCoV-19/USA/WI-MHDL-0662/2021 | EPI_ISL_1652181 |
| hCoV-19/USA/WI-MHDL-0665/2021 | EPI_ISL_1652183 |
| hCoV-19/USA/WI-MHDL-0667/2021 | EPI_ISL_1652185 |
| hCoV-19/USA/WI-MHDL-MU0256/2021 | EPI_ISL_1494740 |
| hCoV-19/USA/WI-MPHL-U1210118/2021 | EPI_ISL_940785 |
| hCoV-19/USA/WI-MHDL-0671/2021 | EPI_ISL_1494753 |
| hCoV-19/USA/WI-MHDL-0680/2021 | EPI_ISL_1652187 |
| hCoV-19/USA/WI-MHDL-0670/2021 | EPI_ISL_1494752 |
| hCoV-19/USA/WI-MHDL-MU0257/2021 | EPI_ISL_1494741 |
| hCoV-19/USA/WI-MHDL-MU0258/2021 | EPI_ISL_1494742 |
| hCoV-19/USA/WI-MHDL-MU0260/2021 | EPI_ISL_1494744 |
| hCoV-19/USA/WI-MHDL-MU0261/2021 | EPI_ISL_1494745 |
| hCoV-19/USA/WI-MHDL-MU0262/2021 | EPI_ISL_1494746 |
| hCoV-19/USA/WI-MHDL-0673/2021 | EPI_ISL_1652188 |

| | |
|---|---|
| hCoV-19/USA/WI-MHDL-MU0263/2021 | EPI_ISL_1494747 |
| hCoV-19/USA/WI-MHDL-U1280177/2021 | EPI_ISL_1493381 |
| hCoV-19/USA/WI-MHDL-U1280187/2021 | EPI_ISL_1493382 |
| hCoV-19/USA/WI-MHDL-0675/2021 | EPI_ISL_1652189 |
| hCoV-19/USA/WI-MHDL-U1280182/2021 | EPI_ISL_1493383 |
| hCoV-19/USA/WI-MHDL-MU0266/2021 | EPI_ISL_1494750 |
| hCoV-19/USA/WI-MHDL-MU0267/2021 | EPI_ISL_1494751 |
| hCoV-19/USA/WI-MHDL-MU0269/2021 | EPI_ISL_1493386 |
| hCoV-19/USA/WI-MHDL-MU0270/2021 | EPI_ISL_1493387 |
| hCoV-19/USA/WI-MHDL-0678/2021 | EPI_ISL_1494737 |
| hCoV-19/USA/WI-MHDL-0681/2021 | EPI_ISL_1494739 |
| hCoV-19/USA/WI-MHDL-MU0279/2021 | EPI_ISL_1262595 |
| hCoV-19/USA/WI-MHDL-MU0281/2021 | EPI_ISL_1262597 |
| hCoV-19/USA/WI-MHDL-U2150085/2021 | EPI_ISL_1262586 |
| hCoV-19/USA/WI-MHDL-MU0284/2021 | EPI_ISL_1262599 |
| hCoV-19/USA/WI-MHDL-MU0285/2021 | EPI_ISL_1262600 |
| hCoV-19/USA/WI-MHDL-MU0286/2021 | EPI_ISL_1262601 |
| hCoV-19/USA/WI-MHDL-U2170020/2021 | EPI_ISL_1262588 |
| hCoV-19/USA/WI-MHDL-U2170019/2021 | EPI_ISL_1262587 |
| hCoV-19/USA/WI-MHDL-MU0288/2021 | EPI_ISL_1262602 |
| hCoV-19/USA/WI-MHDL-MU0290/2021 | EPI_ISL_1262604 |
| hCoV-19/USA/WI-MHDL-U2240053/2021 | EPI_ISL_1262593 |
| hCoV-19/USA/WI-MHDL-U2240049/2021 | EPI_ISL_1262591 |
| hCoV-19/USA/WI-MHDL-MU0291/2021 | EPI_ISL_1262605 |
| hCoV-19/USA/WI-MHDL-MU0292/2021 | EPI_ISL_1262606 |
| hCoV-19/USA/WI-MHDL-MU0293/2021 | EPI_ISL_1262607 |
| hCoV-19/USA/WI-MHDL-MU0298/2021 | EPI_ISL_1494732 |
| hCoV-19/USA/WI-MHDL-MU0299/2021 | EPI_ISL_1494733 |
| hCoV-19/USA/WI-MHDL-MU0300/2021 | EPI_ISL_1494734 |
| hCoV-19/USA/WI-MHDL-0684/2021 | EPI_ISL_1652190 |
| hCoV-19/USA/WI-MHDL-MU0304/2021 | EPI_ISL_1652174 |
| hCoV-19/USA/WI-MHDL-MU0305/2021 | EPI_ISL_1652175 |
| hCoV-19/USA/WI-MHDL-MU0306/2021 | EPI_ISL_1652176 |
| hCoV-19/USA/WI-MHDL-MU0308/2021 | EPI_ISL_1652154 |
| hCoV-19/USA/WI-MHDL-0688/2021 | EPI_ISL_1652161 |
| hCoV-19/USA/WI-MHDL-0691/2021 | EPI_ISL_1592231 |
| hCoV-19/USA/WI-MHDL-0693/2021 | EPI_ISL_1592232 |
| hCoV-19/USA/WI-MHDL-0695/2021 | EPI_ISL_1592234 |
| hCoV-19/USA/WI-MHDL-0700/2021 | EPI_ISL_1592239 |
| hCoV-19/USA/WI-MHDL-0699/2021 | EPI_ISL_1592238 |
| hCoV-19/USA/WI-MHDL-0709/2021 | EPI_ISL_1608148 |
| hCoV-19/USA/WI-MHDL-006155084/2021 | EPI_ISL_2232209 |
| hCoV-19/USA/WI-MHDL-0668/2021 | EPI_ISL_1652186 |

| | |
|---|---|
| hCoV-19/USA/WI-MHDL-0679/2021 | EPI_ISL_1494738 |
| hCoV-19/USA/WI-MHDL-MU0282/2021 | EPI_ISL_1262598 |
| hCoV-19/USA/WI-MHDL-0661/2021 | EPI_ISL_1652180 |
| hCoV-19/USA/WI-MHDL-0666/2021 | EPI_ISL_1652184 |
| hCoV-19/USA/WI-MHDL-0677/2021 | EPI_ISL_1494736 |
| hCoV-19/USA/WI-MHDL-MU0280/2021 | EPI_ISL_1262596 |
| hCoV-19/USA/WI-MHDL-U2240050/2021 | EPI_ISL_1262592 |
| hCoV-19/USA/WI-MHDL-MU0301/2021 | EPI_ISL_1494735 |
| hCoV-19/USA/WI-MHDL-U3120014/2021 | EPI_ISL_1652153 |
| hCoV-19/USA/WI-MHDL-MU0310/2021 | EPI_ISL_1652156 |
| hCoV-19/USA/WI-MHDL-0697/2021 | EPI_ISL_1592236 |
| hCoV-19/USA/WI-MHDL-0696/2021 | EPI_ISL_1592235 |
| hCoV-19/USA/WI-MHDL-0694/2021 | EPI_ISL_1592233 |
| hCoV-19/USA/WI-MHDL-0710/2021 | EPI_ISL_1608149 |
| hCoV-19/USA/WI-MHDL-0713/2021 | EPI_ISL_1608151 |
| hCoV-19/USA/WI-MHDL-4292108/2021 | EPI_ISL_2179059 |
| hCoV-19/USA/WI-MHDL-4192110/2021 | EPI_ISL_2230797 |
| hCoV-19/USA/WI-MHDL-0685/2021 | EPI_ISL_1652159 |
| hCoV-19/USA/WI-MHDL-0698/2021 | EPI_ISL_1592237 |
| hCoV-19/USA/WI-MHDL-0705/2021 | EPI_ISL_1608146 |
| hCoV-19/USA/WI-MHDL-0717/2021 | EPI_ISL_1608155 |
| hCoV-19/USA/WI-MHDL-MU0255/2020 | EPI_ISL_1652170 |
| hCoV-19/USA/WI-MHDL-991264864/2021 | EPI_ISL_2920274 |
| hCoV-19/USA/WI-MHDL-MU0319/2021 | EPI_ISL_1592245 |
| hCoV-19/USA/WI-MHDL-2021120500/2021 | EPI_ISL_8185126 |
| hCoV-19/USA/WI-MHDL-2022010077/2021 | EPI_ISL_8477776 |
| hCoV-19/USA/WI-MHDL-2022010040/2021 | EPI_ISL_8428595 |
| hCoV-19/USA/WI-MHDL-2022010226/2022 | EPI_ISL_8692436 |
| hCoV-19/USA/WI-MHDL-2022010299/2022 | EPI_ISL_8979658 |
| hCoV-19/USA/WI-MHDL-2022010219/2022 | EPI_ISL_8692429 |
| hCoV-19/USA/WI-MHDL-2022010291/2022 | EPI_ISL_8979644 |
| hCoV-19/USA/WI-MHDL-2022020081/2022 | EPI_ISL_9523296 |
| hCoV-19/USA/WI-MHDL-2022020092/2022 | EPI_ISL_9523307 |
| hCoV-19/USA/WI-MHDL-2022020209/2022 | EPI_ISL_9705646 |
| hCoV-19/USA/WI-MHDL-2022010170/2021 | EPI_ISL_8565366 |
| hCoV-19/USA/WI-MHDL-2022010038/2021 | EPI_ISL_8428594 |
| hCoV-19/USA/WI-MHDL-2022010084/2021 | EPI_ISL_8477781 |
| hCoV-19/USA/WI-MHDL-2022010081/2021 | EPI_ISL_8477779 |
| hCoV-19/USA/WI-MHDL-2022010297/2021 | EPI_ISL_8979636 |
| hCoV-19/USA/WI-MHDL-2022010295/2021 | EPI_ISL_8979676 |
| hCoV-19/USA/WI-MHDL-2022010231/2022 | EPI_ISL_8692440 |
| hCoV-19/USA/WI-MHDL-2022010228/2022 | EPI_ISL_8692438 |
| hCoV-19/USA/WI-MHDL-2022010305/2022 | EPI_ISL_8979649 |

| | |
|---|---|
| hCoV-19/USA/WI-MHDL-2022010307/2022 | EPI_ISL_8979655 |
| hCoV-19/USA/WI-MHDL-2022010300/2022 | EPI_ISL_8979665 |
| hCoV-19/USA/WI-MHDL-2022010218/2022 | EPI_ISL_8692428 |
| hCoV-19/USA/WI-MHDL-2022010117/2022 | EPI_ISL_8594429 |
| hCoV-19/USA/WI-MHDL-2022010216/2022 | EPI_ISL_8692426 |
| hCoV-19/USA/WI-MHDL-2022010315/2022 | EPI_ISL_8979647 |
| hCoV-19/USA/WI-MHDL-2022010257/2022 | EPI_ISL_8979654 |
| hCoV-19/USA/WI-MHDL-2022010320/2022 | EPI_ISL_8979632 |
| hCoV-19/USA/WI-MHDL-2022010313/2022 | EPI_ISL_8979651 |
| hCoV-19/USA/WI-MHDL-2022010309/2022 | EPI_ISL_8979662 |
| hCoV-19/USA/WI-MHDL-2022010312/2022 | EPI_ISL_8979673 |
| hCoV-19/USA/WI-MHDL-2022010323/2022 | EPI_ISL_8979678 |
| hCoV-19/USA/WI-MHDL-2022010326/2022 | EPI_ISL_8979675 |
| hCoV-19/USA/WI-MHDL-2022010356/2022 | EPI_ISL_8730943 |
| hCoV-19/USA/WI-MHDL-2022010340/2022 | EPI_ISL_8730929 |
| hCoV-19/USA/WI-MHDL-2022010546/2022 | EPI_ISL_9090404 |
| hCoV-19/USA/WI-MHDL-2022010548/2022 | EPI_ISL_9090406 |
| hCoV-19/USA/WI-MHDL-2022010549/2022 | EPI_ISL_9090407 |
| hCoV-19/USA/WI-MHDL-2022010547/2022 | EPI_ISL_9090405 |
| hCoV-19/USA/WI-MHDL-2022010552/2022 | EPI_ISL_9090410 |
| hCoV-19/USA/WI-MHDL-2022020067/2022 | EPI_ISL_9523283 |
| hCoV-19/USA/WI-MHDL-2022020068/2022 | EPI_ISL_9523284 |
| hCoV-19/USA/WI-MHDL-2022020069/2022 | EPI_ISL_9523285 |
| hCoV-19/USA/WI-MHDL-2022010561/2022 | EPI_ISL_9196451 |
| hCoV-19/USA/WI-MHDL-2022010563/2022 | EPI_ISL_9196453 |
| hCoV-19/USA/WI-MHDL-2022020082/2022 | EPI_ISL_9523297 |
| hCoV-19/USA/WI-MHDL-2022020073/2022 | EPI_ISL_9523289 |
| hCoV-19/USA/WI-MHDL-2022020071/2022 | EPI_ISL_9523287 |
| hCoV-19/USA/WI-MHDL-2022020072/2022 | EPI_ISL_9523288 |
| hCoV-19/USA/WI-MHDL-2022020036/2022 | EPI_ISL_9523259 |
| hCoV-19/USA/WI-MHDL-2022020093/2022 | EPI_ISL_9523308 |
| hCoV-19/USA/WI-MHDL-2022020094/2022 | EPI_ISL_9523309 |
| hCoV-19/USA/WI-MHDL-2022010726/2022 | EPI_ISL_9397749 |
| hCoV-19/USA/WI-MHDL-2022010725/2022 | EPI_ISL_9397748 |
| hCoV-19/USA/WI-MHDL-2022010722/2022 | EPI_ISL_9397745 |
| hCoV-19/USA/WI-MHDL-2022010723/2022 | EPI_ISL_9397746 |
| hCoV-19/USA/WI-MHDL-2022010724/2022 | EPI_ISL_9397747 |
| hCoV-19/USA/WI-MHDL-2022020130/2022 | EPI_ISL_9471961 |
| hCoV-19/USA/WI-MHDL-2022020129/2022 | EPI_ISL_9471964 |
| hCoV-19/USA/WI-MHDL-2022020128/2022 | EPI_ISL_9473318 |
| hCoV-19/USA/WI-MHDL-2022020132/2022 | EPI_ISL_9471945 |
| hCoV-19/USA/WI-MHDL-2022020133/2022 | EPI_ISL_9471947 |
| hCoV-19/USA/WI-MHDL-2022020185/2022 | EPI_ISL_9705638 |

| | |
|---|---|
| hCoV-19/USA/WI-MHDL-2022020186/2022 | EPI_ISL_9705639 |
| hCoV-19/USA/WI-MHDL-2022020309/2022 | EPI_ISL_9766534 |
| hCoV-19/USA/WI-MHDL-2022020213/2022 | EPI_ISL_9705650 |
| hCoV-19/USA/WI-MHDL-2022020208/2022 | EPI_ISL_9705645 |
| hCoV-19/USA/WI-MHDL-2022020210/2022 | EPI_ISL_9705647 |
| hCoV-19/USA/WI-MHDL-2022020211/2022 | EPI_ISL_9705648 |
| hCoV-19/USA/WI-MHDL-2022020212/2022 | EPI_ISL_9705649 |
| hCoV-19/USA/WI-MHDL-2022020207/2022 | EPI_ISL_9705654 |
| hCoV-19/USA/WI-MHDL-2022020351/2022 | EPI_ISL_9812152 |
| hCoV-19/USA/WI-MHDL-2022020334/2022 | EPI_ISL_9766557 |
| hCoV-19/USA/WI-MHDL-2022020335/2022 | EPI_ISL_9766558 |
| hCoV-19/USA/WI-MHDL-2022020336/2022 | EPI_ISL_9766559 |
| hCoV-19/USA/WI-MHDL-2022020332/2022 | EPI_ISL_9766555 |
| hCoV-19/USA/WI-MHDL-2022020333/2022 | EPI_ISL_9766556 |
| hCoV-19/USA/WI-MHDL-2022020337/2022 | EPI_ISL_9766560 |
| hCoV-19/USA/WI-MHDL-2022020338/2022 | EPI_ISL_9766561 |
| hCoV-19/USA/WI-MHDL-2022020368/2022 | EPI_ISL_9812167 |
| hCoV-19/USA/WI-MHDL-2022020481/2022 | EPI_ISL_10033272 |
| hCoV-19/USA/WI-MHDL-2022020478/2022 | EPI_ISL_10033269 |
| hCoV-19/USA/WI-MHDL-2022020479/2022 | EPI_ISL_10033270 |
| hCoV-19/USA/WI-MHDL-2022020567/2022 | EPI_ISL_10023713 |
| hCoV-19/USA/WI-MHDL-2022020568/2022 | EPI_ISL_10023714 |
| hCoV-19/USA/WI-MHDL-2022020569/2022 | EPI_ISL_10023715 |
| hCoV-19/USA/WI-MHDL-2022020584/2022 | EPI_ISL_10023730 |
| hCoV-19/USA/WI-MHDL-2022020583/2022 | EPI_ISL_10023729 |
| hCoV-19/USA/WI-MHDL-2022030100/2022 | EPI_ISL_11032158 |
| hCoV-19/USA/WI-MHDL-2022030031/2022 | EPI_ISL_10645433 |
| hCoV-19/USA/WI-MHDL-2022030391/2022 | EPI_ISL_11506534 |
| hCoV-19/USA/WI-MHDL-2022030213/2022 | EPI_ISL_11160124 |
| hCoV-19/USA/WI-MHDL-2022030214/2022 | EPI_ISL_11160125 |
| hCoV-19/USA/WI-MHDL-2022030549/2022 | EPI_ISL_11301565 |
| hCoV-19/USA/WI-MHDL-2022030409/2022 | EPI_ISL_11506552 |
| hCoV-19/USA/WI-MHDL-2022030681/2022 | EPI_ISL_12059468 |
| hCoV-19/USA/WI-MHDL-2022030744/2022 | EPI_ISL_11816970 |
| hCoV-19/USA/WI-MHDL-2022040399/2022 | EPI_ISL_12746649 |
| hCoV-19/USA/WI-MHDL-2022040288/2022 | EPI_ISL_12431127 |
| hCoV-19/USA/WI-MHDL-2022010304/2022 | EPI_ISL_8979671 |
| hCoV-19/USA/WI-MHDL-2022010259/2022 | EPI_ISL_8979663 |
| hCoV-19/USA/WI-MHDL-2022020032/2022 | EPI_ISL_9523256 |
| hCoV-19/USA/WI-MHDL-2022020095/2022 | EPI_ISL_9523310 |
| hCoV-19/USA/WI-MHDL-2022010314/2022 | EPI_ISL_8979643 |
| hCoV-19/USA/WI-MHDL-2021120385/2021 | EPI_ISL_7877072 |
| hCoV-19/USA/WI-MHDL-2022010169/2021 | EPI_ISL_8565365 |

| | |
|---|---|
| hCoV-19/USA/WI-MHDL-2022010021/2021 | EPI_ISL_8388867 |
| hCoV-19/USA/WI-MHDL-2022010225/2022 | EPI_ISL_8692435 |
| hCoV-19/USA/WI-MHDL-2022010118/2022 | EPI_ISL_8594430 |
| hCoV-19/USA/WI-MHDL-2022010217/2022 | EPI_ISL_8692427 |
| hCoV-19/USA/WI-MHDL-2022010518/2022 | EPI_ISL_8879387 |
| hCoV-19/USA/WI-MHDL-2022010355/2022 | EPI_ISL_8730942 |
| hCoV-19/USA/WI-MHDL-2022020031/2022 | EPI_ISL_9523255 |
| hCoV-19/USA/WI-MHDL-2022010721/2022 | EPI_ISL_9397744 |
| hCoV-19/USA/WI-MHDL-2022020214/2022 | EPI_ISL_9705651 |
| hCoV-19/USA/WI-MHDL-2022010087/2021 | EPI_ISL_8477784 |
| hCoV-19/USA/WI-MHDL-2022010088/2021 | EPI_ISL_8477785 |
| hCoV-19/USA/WI-MHDL-2022010358/2022 | EPI_ISL_8730945 |
| hCoV-19/USA/WI-MHDL-2022010224/2022 | EPI_ISL_8692434 |
| hCoV-19/USA/WI-MHDL-2022010223/2022 | EPI_ISL_8692433 |
| hCoV-19/USA/WI-MHDL-2022010230/2022 | EPI_ISL_8692439 |
| hCoV-19/USA/WI-MHDL-2022010227/2022 | EPI_ISL_8692437 |
| hCoV-19/USA/WI-MHDL-2022010306/2022 | EPI_ISL_8979619 |
| hCoV-19/USA/WI-MHDL-2022010303/2022 | EPI_ISL_8979657 |
| hCoV-19/USA/WI-MHDL-2022010302/2022 | EPI_ISL_8979660 |
| hCoV-19/USA/WI-MHDL-2022010265/2022 | EPI_ISL_8979664 |
| hCoV-19/USA/WI-MHDL-2022010294/2022 | EPI_ISL_8979624 |
| hCoV-19/USA/WI-MHDL-2022010292/2022 | EPI_ISL_8979631 |
| hCoV-19/USA/WI-MHDL-2022010308/2022 | EPI_ISL_8979633 |
| hCoV-19/USA/WI-MHDL-2022010322/2022 | EPI_ISL_8979667 |
| hCoV-19/USA/WI-MHDL-2022010321/2022 | EPI_ISL_8979653 |
| hCoV-19/USA/WI-MHDL-2022010311/2022 | EPI_ISL_8979666 |
| hCoV-19/USA/WI-MHDL-2022010357/2022 | EPI_ISL_8730944 |
| hCoV-19/USA/WI-MHDL-2022010327/2022 | EPI_ISL_8979630 |
| hCoV-19/USA/WI-MHDL-2022010339/2022 | EPI_ISL_8730928 |
| hCoV-19/USA/WI-MHDL-2022010353/2022 | EPI_ISL_8730940 |
| hCoV-19/USA/WI-MHDL-2022010351/2022 | EPI_ISL_8730939 |
| hCoV-19/USA/WI-MHDL-2022010354/2022 | EPI_ISL_8730941 |
| hCoV-19/USA/WI-MHDL-2022010544/2022 | EPI_ISL_9090402 |
| hCoV-19/USA/WI-MHDL-2022010564/2022 | EPI_ISL_9196454 |
| hCoV-19/USA/WI-MHDL-2022020033/2022 | EPI_ISL_9523257 |
| hCoV-19/USA/WI-MHDL-2022020131/2022 | EPI_ISL_9471956 |
| hCoV-19/USA/WI-MHDL-2022020582/2022 | EPI_ISL_10023728 |
| hCoV-19/USA/WI-MHDL-2022010180/2021 | EPI_ISL_8565373 |
| hCoV-19/USA/WI-MHDL-2022010036/2021 | EPI_ISL_8428592 |
| hCoV-19/USA/WI-MHDL-2022010083/2021 | EPI_ISL_8477780 |
| hCoV-19/USA/WI-MHDL-2022010222/2022 | EPI_ISL_8692432 |
| hCoV-19/USA/WI-MHDL-2022010119/2022 | EPI_ISL_8594431 |
| hCoV-19/USA/WI-MHDL-2022010120/2022 | EPI_ISL_8594432 |

| | |
|---|---|
| hCoV-19/USA/WI-MHDL-2022010325/2022 | EPI_ISL_8979627 |
| hCoV-19/USA/WI-MHDL-2022010342/2022 | EPI_ISL_8730931 |
| hCoV-19/USA/WI-MHDL-2022010551/2022 | EPI_ISL_9090409 |
| hCoV-19/USA/WI-MHDL-2022020064/2022 | EPI_ISL_9523279 |
| hCoV-19/USA/WI-MHDL-2022020066/2022 | EPI_ISL_9523281 |
| hCoV-19/USA/WI-MHDL-2022020070/2022 | EPI_ISL_9523286 |
| hCoV-19/USA/WI-MHDL-2022020035/2022 | EPI_ISL_9523258 |
| hCoV-19/USA/WI-MHDL-2022020206/2022 | EPI_ISL_9705644 |
| hCoV-19/USA/WI-MHDL-2021120363/2021 | EPI_ISL_7744413 |
| hCoV-19/USA/WI-MHDL-2021120368/2021 | EPI_ISL_7744418 |
| hCoV-19/USA/WI-MHDL-2021120390/2021 | EPI_ISL_7877075 |
| hCoV-19/USA/WI-MHDL-2021120392/2021 | EPI_ISL_7877076 |
| hCoV-19/USA/WI-MHDL-2021120502/2021 | EPI_ISL_8185128 |
| hCoV-19/USA/WI-MHDL-2022010179/2021 | EPI_ISL_8593660 |
| hCoV-19/USA/WI-MHDL-2022010181/2021 | EPI_ISL_8593661 |
| hCoV-19/USA/WI-MHDL-2022010076/2021 | EPI_ISL_8483086 |
| hCoV-19/USA/WI-MHDL-2022010078/2021 | EPI_ISL_8477777 |
| hCoV-19/USA/WI-MHDL-2022010079/2021 | EPI_ISL_8477778 |
| hCoV-19/USA/WI-MHDL-2022010034/2021 | EPI_ISL_8428591 |
| hCoV-19/USA/WI-MHDL-2022010033/2021 | EPI_ISL_8428590 |
| hCoV-19/USA/WI-MHDL-2022010032/2021 | EPI_ISL_8428589 |
| hCoV-19/USA/WI-MHDL-2022010039/2021 | EPI_ISL_8428813 |
| hCoV-19/USA/WI-MHDL-2022010085/2021 | EPI_ISL_8477782 |
| hCoV-19/USA/WI-MHDL-2022010186/2021 | EPI_ISL_8565375 |
| hCoV-19/USA/WI-MHDL-2022010037/2021 | EPI_ISL_8428593 |
| hCoV-19/USA/WI-MHDL-2022010082/2021 | EPI_ISL_8483087 |
| hCoV-19/USA/WI-MHDL-2022010086/2021 | EPI_ISL_8477783 |
| hCoV-19/USA/WI-MHDL-2022010090/2021 | EPI_ISL_8477786 |
| hCoV-19/USA/WI-MHDL-2022010296/2021 | EPI_ISL_8979661 |
| hCoV-19/USA/WI-MHDL-2022010298/2022 | EPI_ISL_8979617 |
| hCoV-19/USA/WI-MHDL-2022010264/2022 | EPI_ISL_8979634 |
| hCoV-19/USA/WI-MHDL-2022010301/2022 | EPI_ISL_8979639 |
| hCoV-19/USA/WI-MHDL-2022010220/2022 | EPI_ISL_8692430 |
| hCoV-19/USA/WI-MHDL-2022010262/2022 | EPI_ISL_8979629 |
| hCoV-19/USA/WI-MHDL-2022010256/2022 | EPI_ISL_8979614 |
| hCoV-19/USA/WI-MHDL-2022010310/2022 | EPI_ISL_8979638 |
| hCoV-19/USA/WI-MHDL-2022010328/2022 | EPI_ISL_8979670 |
| hCoV-19/USA/WI-MHDL-2022010341/2022 | EPI_ISL_8730930 |
| hCoV-19/USA/WI-MHDL-2022010543/2022 | EPI_ISL_9090401 |
| hCoV-19/USA/WI-MHDL-2022010545/2022 | EPI_ISL_9090403 |
| hCoV-19/USA/WI-MHDL-2022010550/2022 | EPI_ISL_9090408 |
| hCoV-19/USA/WI-MHDL-2022020065/2022 | EPI_ISL_9523280 |
| hCoV-19/USA/WI-MHDL-2022020096/2022 | EPI_ISL_9523311 |

| | |
|---|---|
| hCoV-19/USA/WI-MHDL-2022020134/2022 | EPI_ISL_9471965 |
| hCoV-19/USA/WI-MHDL-2022020127/2022 | EPI_ISL_9471955 |
| hCoV-19/USA/WI-MHDL-2022020480/2022 | EPI_ISL_10033271 |
| hCoV-19/USA/WI-MHDL-2022030550/2022 | EPI_ISL_11301566 |
| hCoV-19/USA/WI-MHDL-2022030565/2022 | EPI_ISL_11360117 |
| hCoV-19/USA/WI-MHDL-2022030566/2022 | EPI_ISL_11360118 |
| hCoV-19/USA/WI-MHDL-2022030410/2022 | EPI_ISL_11506553 |
| hCoV-19/USA/WI-MHDL-2022040400/2022 | EPI_ISL_12746650 |
| hCoV-19/USA/WI-MHDL-2022040283/2022 | EPI_ISL_12431122 |
| hCoV-19/USA/WI-MHDL-2022040284/2022 | EPI_ISL_12431123 |
| hCoV-19/USA/WI-MHDL-2022050015/2022 | EPI_ISL_12713286 |
| hCoV-19/USA/WI-MHDL-2022010562/2022 | EPI_ISL_9196452 |
| hCoV-19/USA/WI-MHDL-2022020084/2022 | EPI_ISL_9523299 |
| hCoV-19/USA/WI-MHDL-2022040285/2022 | EPI_ISL_12431124 |
| hCoV-19/USA/WI-MHDL-2022040287/2022 | EPI_ISL_12431126 |
| hCoV-19/USA/WI-MHDL-2022040291/2022 | EPI_ISL_12431130 |
| hCoV-19/USA/WI-MHDL-2022050014/2022 | EPI_ISL_12713285 |
| hCoV-19/USA/WI-MHDL-2022030746/2022 | EPI_ISL_11816972 |
| hCoV-19/USA/WI-MHDL-2022040401/2022 | EPI_ISL_12746651 |
| hCoV-19/USA/WI-MHDL-2022050013/2022 | EPI_ISL_12713284 |
| hCoV-19/USA/WI-MHDL-2022030745/2022 | EPI_ISL_11816971 |
| hCoV-19/USA/WI-MHDL-2022040286/2022 | EPI_ISL_12431125 |
| hCoV-19/USA/WI-MHDL-MU0259/2021 | EPI_ISL_1494743 |
| hCoV-19/USA/WI-MHDL-MU0264/2021 | EPI_ISL_1494748 |
